# Machine Learning-based Prediction of Preterm Birth Using Genetic Data

**DOI:** 10.64898/2026.06.24.26356330

**Authors:** Hedvig Sundelin, Bo Jacobsson, Karin Ytterberg, Pol Sole-Navais, Julius Juodakis

## Abstract

The leading cause of mortality and morbidity in children under the age of 5 is preterm birth. The timing of birth is influenced by both genetic and environmental factors, but the underlying mechanisms remain poorly understood, making its prediction difficult. In this study, we investigated the potential of using machine learning models to predict preterm birth based on genetic data from the Norwegian Mother, Father and Child Cohort Study (MoBa). We trained and evaluated several classification algorithms on individual-level genetic data from over 15,000 mothers and children. Our results indicate that the predictive capacity of maternal gestational duration-associated loci for preterm birth is limited, with the highest AUC values around 0.57. Additionally, incorporating more SNPs within the associated loci did not improve prediction performance. As expected, the contribution of the maternal genome to preterm birth prediction was found to be larger than that of the fetal genome. Overall, our findings suggest that while genetic testing provides some information about an individual’s risk for preterm birth, further research incorporating additional factors is necessary to enhance predictability.

## 1 Introduction

Preterm Birth (PTB) refers to delivery before 37 completed weeks of gestation. It is the leading cause of death in children under the age of 5[1]. Additionally, surviving children risk having their long-term health affected in several ways, from respiratory problems to cerebral palsy, bronchopulmonary dysplasia, epilepsy and hyperbilirubinemia [2]–[5]. The risks associated with PTB could be reduced through close monitoring and appropriate medical care but would require better stratification of women at risk [6].

Despite knowledge of several factors that increase the risk of PTB, including high blood pressure, diabetes, stress, or infections [7], the cause of most PTB remains unknown [8]. Several studies have shown that genetic factors play a role in pregnancy duration, with pedigree-based heritability estimates of around 20-30% [9]–[12]. A recent genome-wide association study identified 25 variants in the mother’s genome associated with either PTB, gestational duration or both [13]. However, the effect sizes of these variants are relatively small and do not explain the total hereditary proportion of variation in the gestational duration. In fact, only 2.2% of the variance in gestational duration was explained after using a linear model of approximately 1 million variants [13].

The simple models currently used to predict gestational duration or other traits are based on additive effects of genetic variants. More complex models, involving gene-gene interactions or non-additive effects have largely been overlooked. One study that utilized machine learning models for predicting PTB was conducted in 2018. The genotyped data consisted of 1 527 mothers of African or Haitian descent, of which around 40% were classified as having had preterm deliveries [14]. The authors report that using 5000 genetic variants provide an almost perfect prediction accuracy when used to train their network model.

This project aims to further investigate the viability in using machine learning models to predict PTB and whether this could be used in identifying high risk individuals at an early stage. Several different methods for classification are trained and tested on individual-level genetic data from >15 000 mothers and children in the Norwegian Mother, Father and Child Cohort Study (MoBa).

## 2 Materials and Methods

### Ethics

This project’s analyses have received approval from the Swedish (Etikprövningsmyndigheten, decision Dnr 2022-03248-01) and Norwegian (Regionale komiteer for medisinsk og helsefaglig forskningsetikk sør-øst project 2015/2425) institutional review boards. All research activities align with the principles outlined in the Helsinki declaration. Genetic data collection was performed with informed consent from participants.

### Data Source

MoBa is a population-based pregnancy cohort study conducted by the Norwegian Institute of Public Health [15]. Participants were recruited from all over Norway from 1999–2008. The cohort includes approximately 114 500 children, 95 200 mothers, and 75 200 fathers. The data used in this study is version 12 of the quality-assured data files released by MoBa. It includes genotype data, questionnaires filled out by the parents, and linked records from Medical Birth Registry of Norway (MBRN) [16], a national health registry containing information about all births in Norway.

The present study used a subset of this cohort: pregnancy outcomes, maternal and fetal genotypes from ∼31 000 parent-offspring trios that were genotyped over 2012–2018. Full description of genotyping, phasing and quality control can be found in a previously published study [17].

### Sample selection and phenotyping

Genotyped samples were restricted to singleton, live-birth pregnancies with complete birth registry data, at least one answered MoBa questionnaire, and individuals alive at the time of genotyping. Additionally, we excluded pregnancies involving IVF, maternal diabetes, extreme outlier gestational duration, and children with an APGAR score of 0 at both 1 and 5 minutes. Finally, only deliveries with spontaneous onset were included and one random pregnancy was retained for mothers with repeated pregnancies in the cohort. By implementing these inclusion criteria, the study minimizes potential confounding (i.e. when ethnicity affects both genetics and the outcome), reducing the risk of identified associations between genotypes and outcomes being influenced by population stratification.

We analyzed a total of 16 301 pregnancies, each linked to both maternal and fetal genotyped data. Only pregnancies with data from both genomes were considered, ensuring accurate comparisons of their respective predictability.

Gestational duration, in days (SVLEN_DG [18]), was used to categorize PTB as follows:

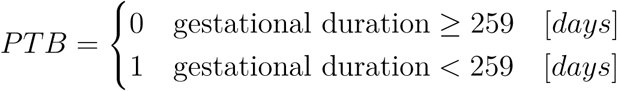

This binary classification resulted in 15 882 control samples and 419 case samples.

### Model training and prediction

We tested 96 combinations of feature sets (Table 1) and classification models (Table 2) to create predictions of the target variable, PTB.

**Table 1:** Feature sets, each set of features come in three different versions.

| Name | Reduction | Selection | Features |
| --- | --- | --- | --- |
| Full | GWAS | No |  |
| - Maternal |  |  | 7 833 |
| - Fetal |  |  | 7 833 |
| - Combined |  |  | 15 660 |
| Top29 | GWAS | $p\text{-value} < 5e^{-8}$ | |
| - Maternal |  |  | 29 |
| - Fetal |  |  | 29 |
| - Combined |  |  | 58 |
| Top5 | GWAS | min p-value |  |
| - Maternal |  |  | 5 |
| - Fetal |  |  | 5 |
| - Combined |  |  | 10 |
| Selected | GWAS + f-anova | top k |  |
| - Maternal |  |  | 100 |
| - Fetal |  |  | 100 |
| - Combined |  |  | 200 |

**Table 2:** Classification models and properties.

| Classifier | Parametric<sup>1</sup> | Assumptions | Advantages & Disadvantages |
| --- | --- | --- | --- |
| Logistic regression | Yes | Linear decision boundary, assumes independent features | Simple, fast, probability estimates.<br>Not suitable for complex relationships |
| Linear Discriminant Analysis (LDA) | Yes | Assumes normal distribution and equal covariance matrices | Robust to correlated features<br>Sensitive to outliers |
| Quadratic Discriminant Analysis (QDA) | Yes | Assumes normal distribution, allows different covariance matrices | Handles multi-class problems<br>Sensitive to outliers |
| Support vector | No | None | Robust to overfitting, handles high dimensional data<br>Requires good choice of kernel, not suitable for large datasets |
| Random forest | No | None | Robust to overfitting, handles high dimensional data, feature importance<br>Not interpretable, can be slow |
| Bernoulli Naive Bayes (B. Naive Bayes) | Yes | Assumes binary, independent features | Fast, works well with high dimensions<br>Independence assumption may not hold |
| k-Nearest neighbors | No | None | Simple, no training time<br>Slow prediction time, sensitive to irrelevant features |
| Neural network | No | None | Can model complex relationships, handles high dimensional data<br>Requires a lot of data, sensitive to feature scaling, non-interpretable |

<sup>1</sup>**Parametric:** Indicates whether the classifier makes specific assumptions about data distribution with a fixed number of parameters.

The model development flowchart is shown in Fig. 1; for each configuration, we used a stratified 5-fold cross-validation split to evaluate model performance on the selected feature set. In each fold, data was extracted and preprocessed according to the selected configuration and divided into training and test data. The model was then trained in two steps, starting with hyperparameter tuning. Using the tuned hyperparameters, the model was fitted on 20 different versions of randomly undersampled training data and used, on the test set, to predict the outcome, PTB. We then calculated the average prediction over all iterations and evaluated performance based on how well the prediction matched the true values. The resulting metrics for each fold were thereafter combined to obtain an average over all folds.

**Figure 1:**
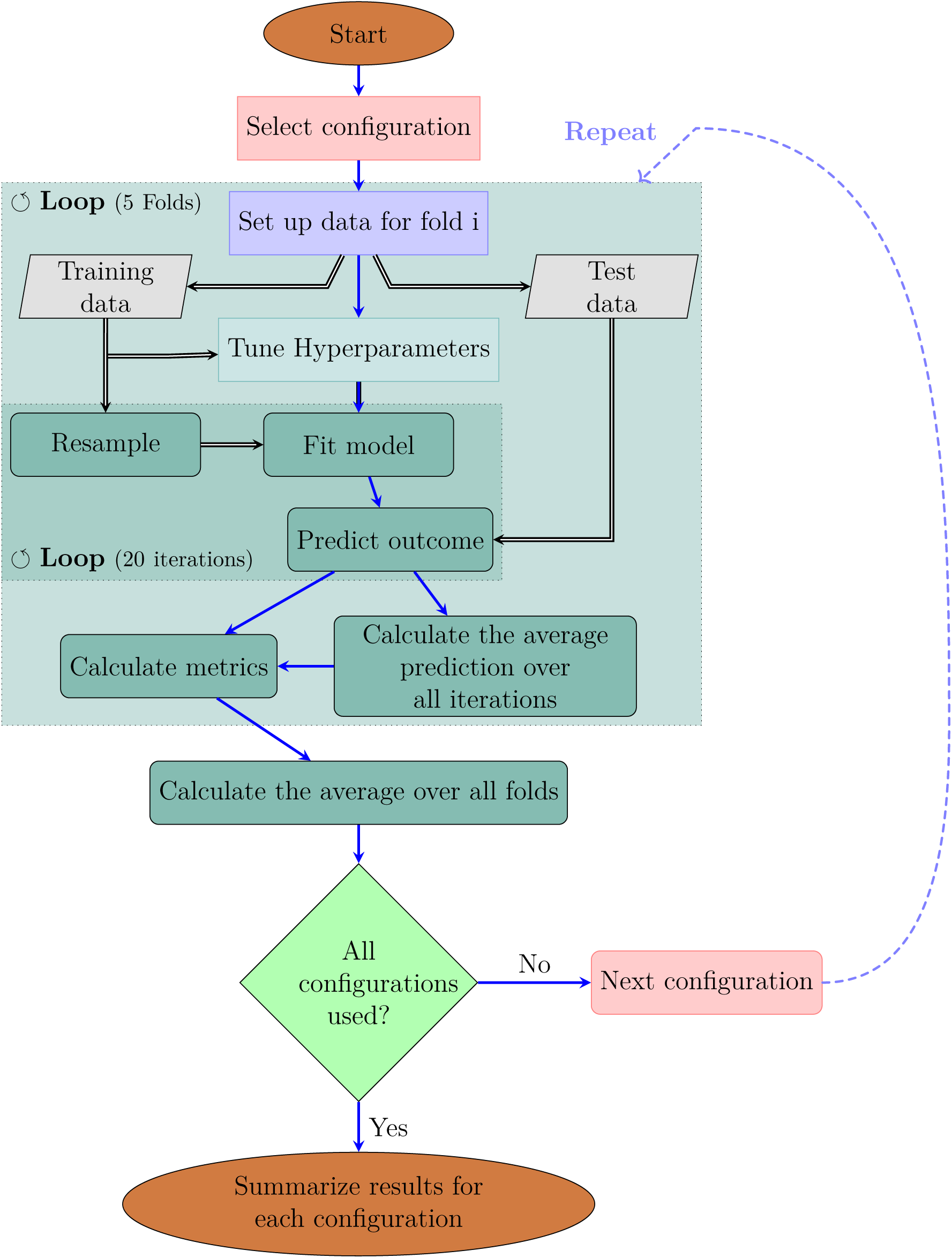
Flow diagram over the process of model training and evaluation.

**Figure 2:**
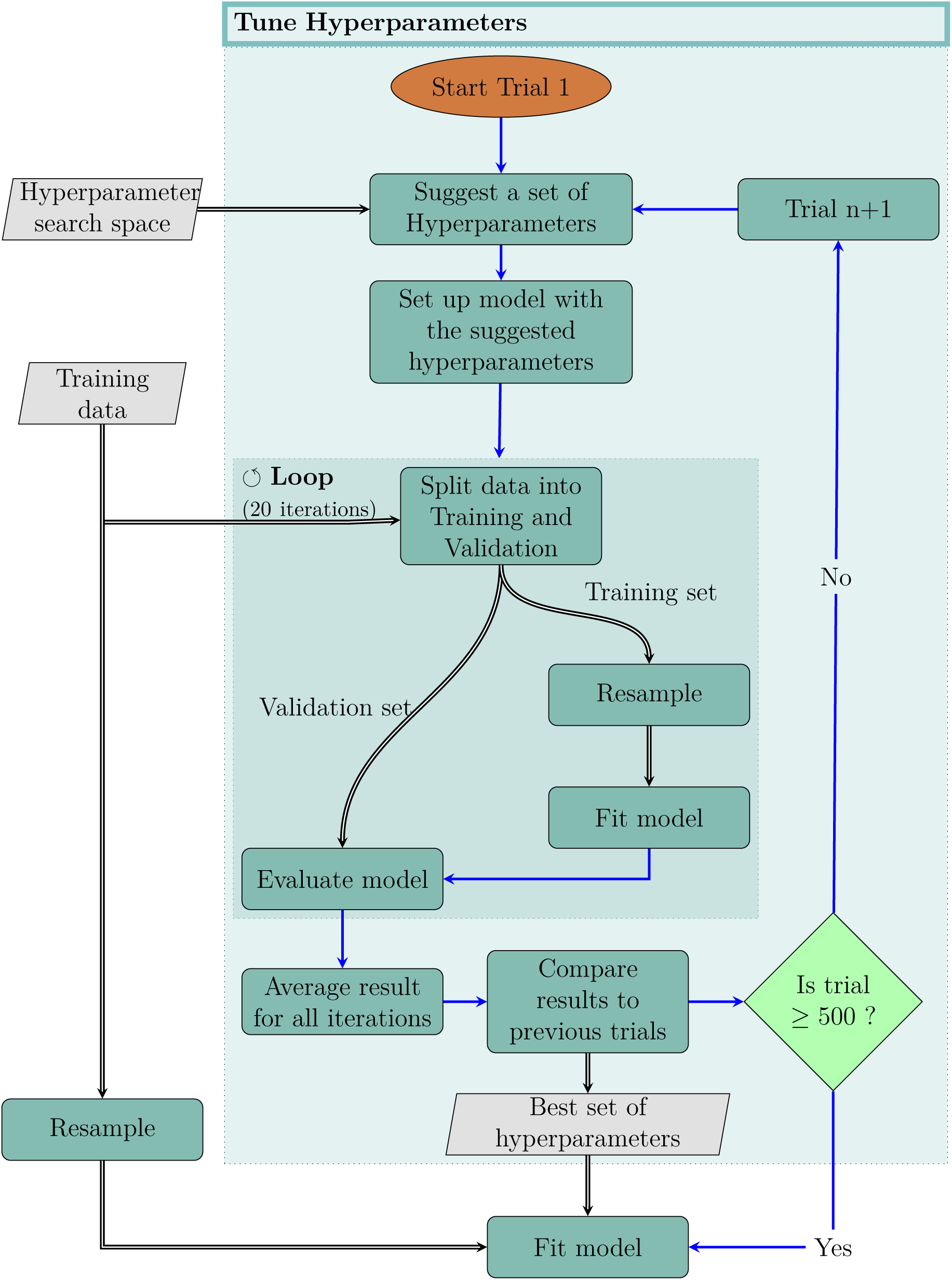
Flow diagram expanding on the hyperparameter tuning process

#### Feature sets

Given that using all imputed genetic variants would increase the noise-to-signal ratio, we based this study on a subset of genetic regions that were previously identified in the largest Genome-wide association study (GWAS) to date [13]. We used all imputed SNPs within 50 kb from the genome-wide significant SNPs in the previous GWAS [13], and applied clumping and thresholding using PLINK 1.9 [19], [20], with a step size of 5 variants and an *r*^2^ threshold of 0.3.

The resulting subset of features consisted of 7 833 Single Nucleotide Polymorphisms (SNPs) and is henceforth referred to as the “full” set, Table 1. From the “full” set three feature subsets of varying size were created. The feature set “top5” contain the five SNPs with the lowest p-value, while “top29” contain all SNPs with a p-value below 5e-8 (n = 29). For the last feature set, we implemented an additional feature reduction method. We looked at which features contained the highest ratio of between-group variability and within-group variability.SelectKBest[21] was implemented to extract the, k=100, highest scoring features, in each set of training data, according to the ANOVA f-score (Eq. (1)).

#### Classification Models

The models and algorithms we used employ different approaches to classification, ranging from linear models, such as logistic regression, to non-parametric models, such as k-Nearest Neighbors, to ensemble models, such as Random Forest. Each algorithm has its strengths and weaknesses, and the choice of algorithm depends on the specific problem and data available. For the problem at hand, there was no determined ‘best choice’, which is why we decided to look at several different models. In Table 2, we list the eight different machine learning models that we used in this project, together with some of their properties.

#### Hyperparameter tuning

It is worth noting that the performance of a machine learning model depends not only on the choice of algorithm but also on the quality and quantity of the data used for training and testing [22], as well as on hyperparameters. The latter are set before training and control the model’s behavior, which can significantly impact its performance[23]. Hyperparameter tuning was done with Optuna, a hyperparameter optimization framework [24]. The tuning operation in Optuna is called a ‘Study’. Each study included 500 trials, each with 20 iterations of training the model with a set of hyperparameters sampled by Optunas ‘TPESampling’ using a Tree-structured Parzen Estimator (TPE) algorithm [25] and evaluating it on a validation set.

The hyperparameters that provided the highest average Area Under the Curve (AUC) on the validation set over the 20 iterations were saved and used to set up the model before fitting it on the full training set.

#### Resampling

The available data is unbalanced, resampling the data (either under or over) makes it easier for classification models to detect useful patterns. Oversampling is significantly more time-consuming and early testing showed a better response to undersampled data, Appendix A. The resampling process was therefore done by random undersampling [26]. This is the most simple undersampling method, chosen since more complex methods did not show any indications of improving results, Fig. S3.

### Evaluation

The evaluation of the models was conducted using various metrics to assess their performance in predicting preterm delivery. The Matthews Correlation Coefficient (MCC) (Eq. (5)), which takes all elements in the confusion matrix into account[27], the AUC (Eq. (2)), and Odds Ratio (OR) (Eq. (6)) was calculated for each model across different feature sets, providing a good view of their classification capabilities. In addition to these, Sensitivity and Specificity were calculated to provide a comprehensive view of the balance between classifications.

#### Model Assessment

Model fitting was repeated 20 times, each time on a new random undersampling of the training data. The classification of each test sample was based on calculating the probabilities of belonging to either class. The predicted probabilities were first used to calculate performance metrics for each repeat individually. In addition to this, we created new predicted probabilities by combining the predictions from all repeats. This combined prediction was also used to calculate performance metrics. By looking at the difference between metrics for the combined prediction and the average metrics for all repeats we can see whether the variability is due to differences between folds or if the model itself is unstable.

A previous study [13] built a Polygenic Risk Score (PRS) for gestational duration to explore the potential utility of their identified SNPs. Using this score for predicting preterm delivery provides a median AUC of 0.595 (mean: 0.582). However, the score is based on the weights from 1 million common SNPs and partially trained on some of the samples used in this study. The prediction could therefore be biased and overfitted to some degree.

## 3 Results

### Gestational duration-associated loci do not contribute to preterm birth prediction beyond maternal top SNPs

We first assessed the predictive capacity of maternal gestational duration-related loci regarding preterm delivery. The examination of the top 29 SNPs in the maternal genome showed that the Naive Bayes algorithm attained the highest mean and median AUC (0.567, 0.570 respectively), the highest OR (1.499) and alongside the Support vector the highest MCC, as illustrated in Table 3. Except for the k-Nearest neighbor model, the other algorithms demonstrated comparable performance, with median AUC values fluctuating between 0.54 and 0.56. Given that our models incorporated gene-gene interactions and non-additive effects, these effects appear to be meager in the gestational duration-associated loci.

**Table 3:** Metrics by model on the Top29 subset in the maternal genome.

| Model | AUC |  |  | OR |  | Sens | Spec | MCC |
| --- | --- | --- | --- | --- | --- | --- | --- | --- |
|  | mean | median | 95% CI | mean | 95% CI | mean | mean | mean |
| B Naive Bayes | 0.567 | 0.570 | (0.56, 0.57) | 1.499 | (1.44, 1.56) | 0.537 | 0.558 | 0.030 |
| LDA | 0.556 | 0.555 | (0.55, 0.56) | 1.382 | (1.34, 1.43) | 0.513 | 0.564 | 0.025 |
| Log. Regression | 0.556 | 0.556 | (0.55, 0.56) | 1.418 | (1.37, 1.47) | 0.531 | 0.551 | 0.026 |
| Neural Network | 0.537 | 0.537 | (0.53, 0.54) | 1.275 | (1.22, 1.33) | 0.530 | 0.518 | 0.016 |
| QDA | 0.552 | 0.553 | (0.55, 0.56) | 1.353 | (1.31, 1.4) | 0.507 | 0.565 | 0.023 |
| Random Forest | 0.564 | 0.563 | (0.56, 0.57) | 1.458 | (1.41, 1.51) | 0.543 | 0.547 | 0.029 |
| Support Vector | 0.561 | 0.559 | (0.56, 0.57) | 1.492 | (1.44, 1.55) | 0.543 | 0.552 | 0.030 |
| k-Nearest Neighbor | 0.523 | 0.526 | (0.52, 0.53) | 1.134 | (1.07, 1.19) | 0.321 | 0.701 | 0.008 |

Fig. 3 shows the test and training AUCs for different models on the top 29 SNPs in the maternal genome. The training AUCs for the Random forest and Neural network models are substantially higher than the test AUCs, which suggests that these models may be overfitting to the training data.

**Figure 3:**
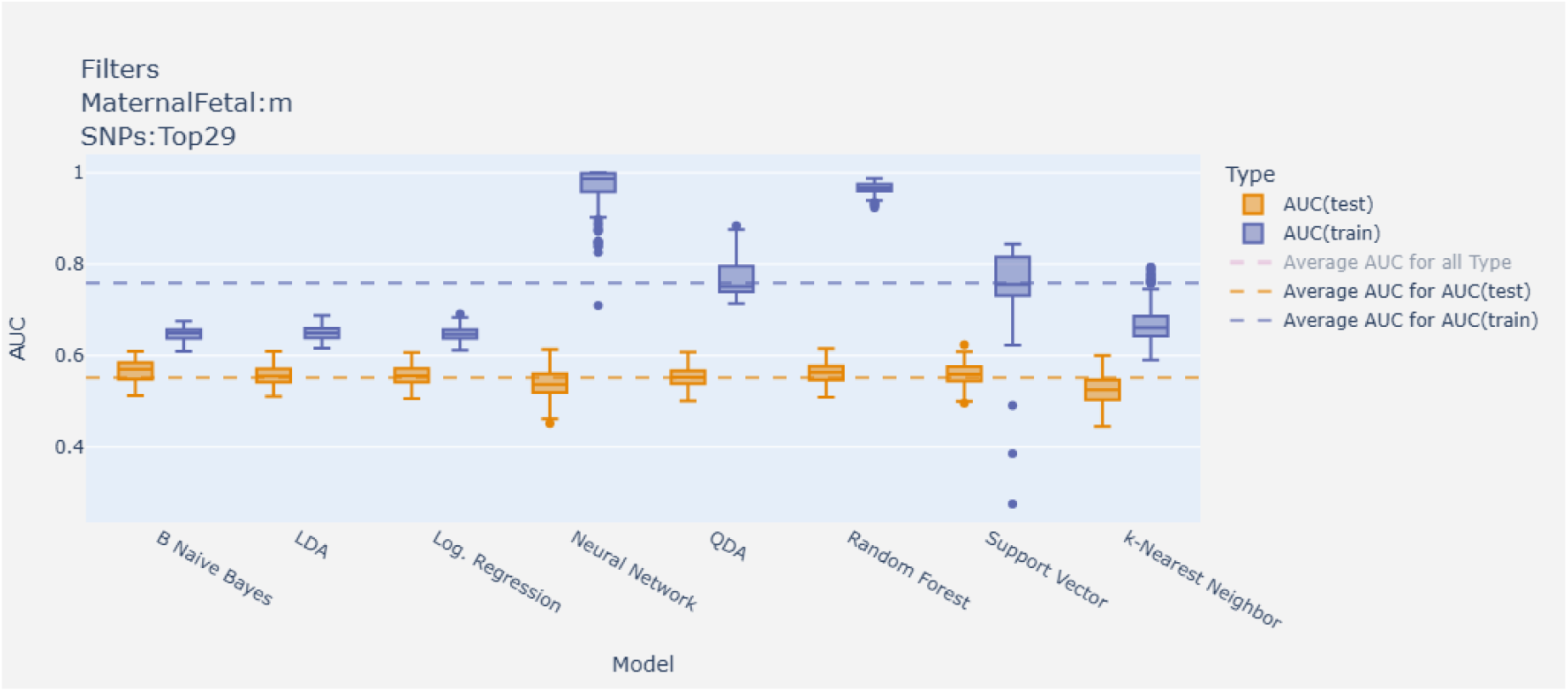
Test and training results (AUC) by model on the top 29 SNPs in the maternal genome.

As seen in Fig. 4, model performance did not increase linearly with the number of features considered in the maternal genome. The highest average AUC (0.55) was obtained with the top 29 SNPs, closely followed by the Top5 subset (Average AUC: 0.54). These results indicate that additional SNPs within the associated loci do not contribute to preterm birth prediction beyond the effect of the top 29 SNPs. Limiting the SNPs to only the top 5 decreased predictive performance to some extent, indicating that there are informative SNPs beyond these.

**Figure 4:**
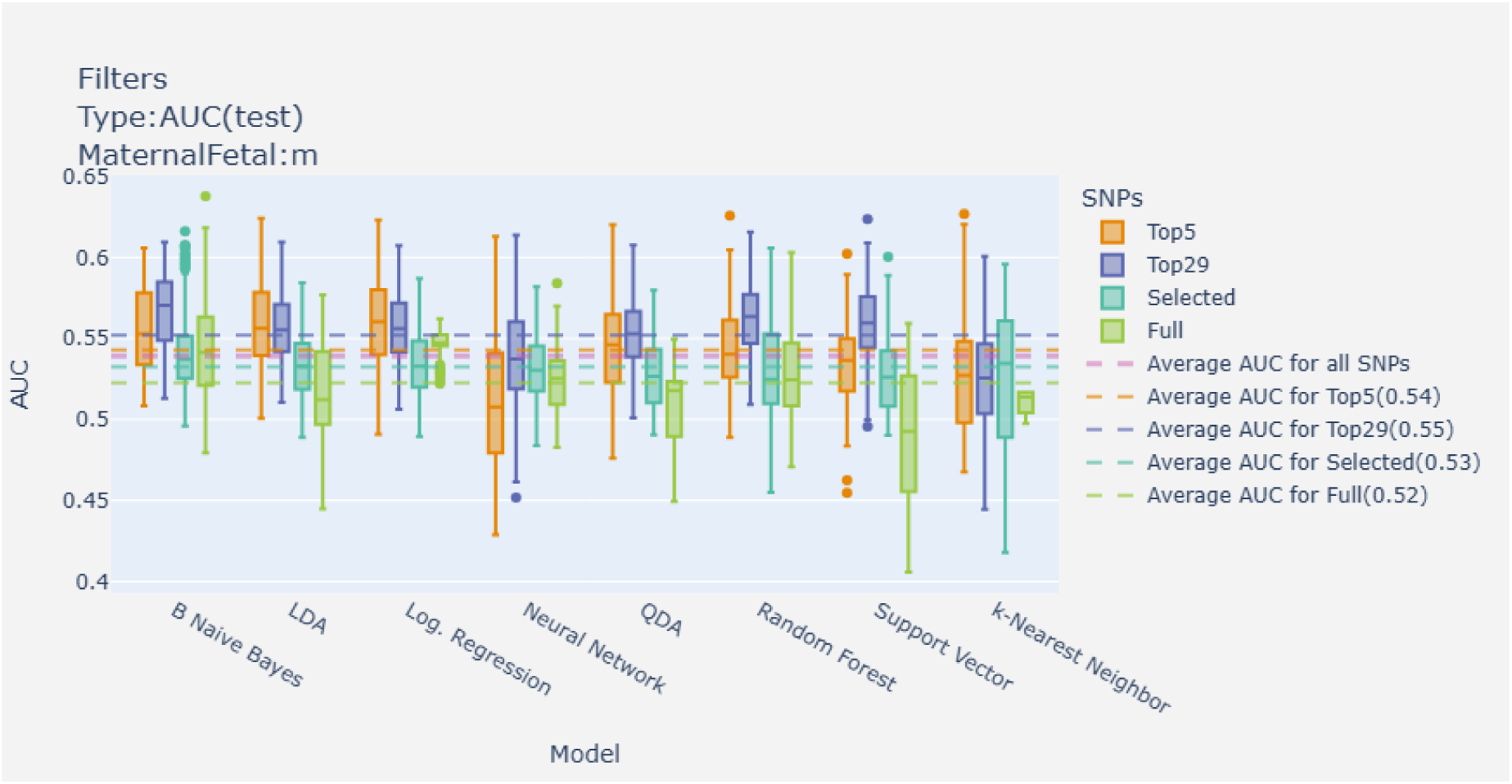
AUC by model on different sets of SNPs in the maternal genome.

### Feature selection gives weak predictions but reverses the maternal-fetal performance pattern

The Selected subset, based on the SNPs creating the highest ratio of variance between groups and variance within groups, performed poorly on average for the maternal genome, Fig. 4. If we look at the variants selected in the maternal genome, Table S10, only four SNPs have a p-value below 5e-8 and only 12 variants are selected for all five folds. In the fetal genome only one SNP have a p-value below 5e-8 while 14 variants are selected for all five folds. While results are highly variable and not particularly accurate,Fig. S6, it is noteworthy that the average AUC in the Selected subset is lowest for the maternal genome.

### The contribution of the maternal genome is larger than that of the fetal one

In Fig. 5 we can see how the models performed on the top 29 SNPS from different genomes, (maternal, fetal and combined). Using the top 29 SNPs from the fetal genome, we obtain very limited predictive accuracy (Average AUC: 0.51). The loci used in this study were discovered by analysing the maternal genome and the contribution of these loci in the fetal genome seem to be minimal at best. Combining the maternal and fetal genome in the same model does not improve predictions in general. Although there is one model, k-Nearest neighbor, which performs better on the combined genome than on the maternal genome alone this model is outperformed by others.

**Figure 5:**
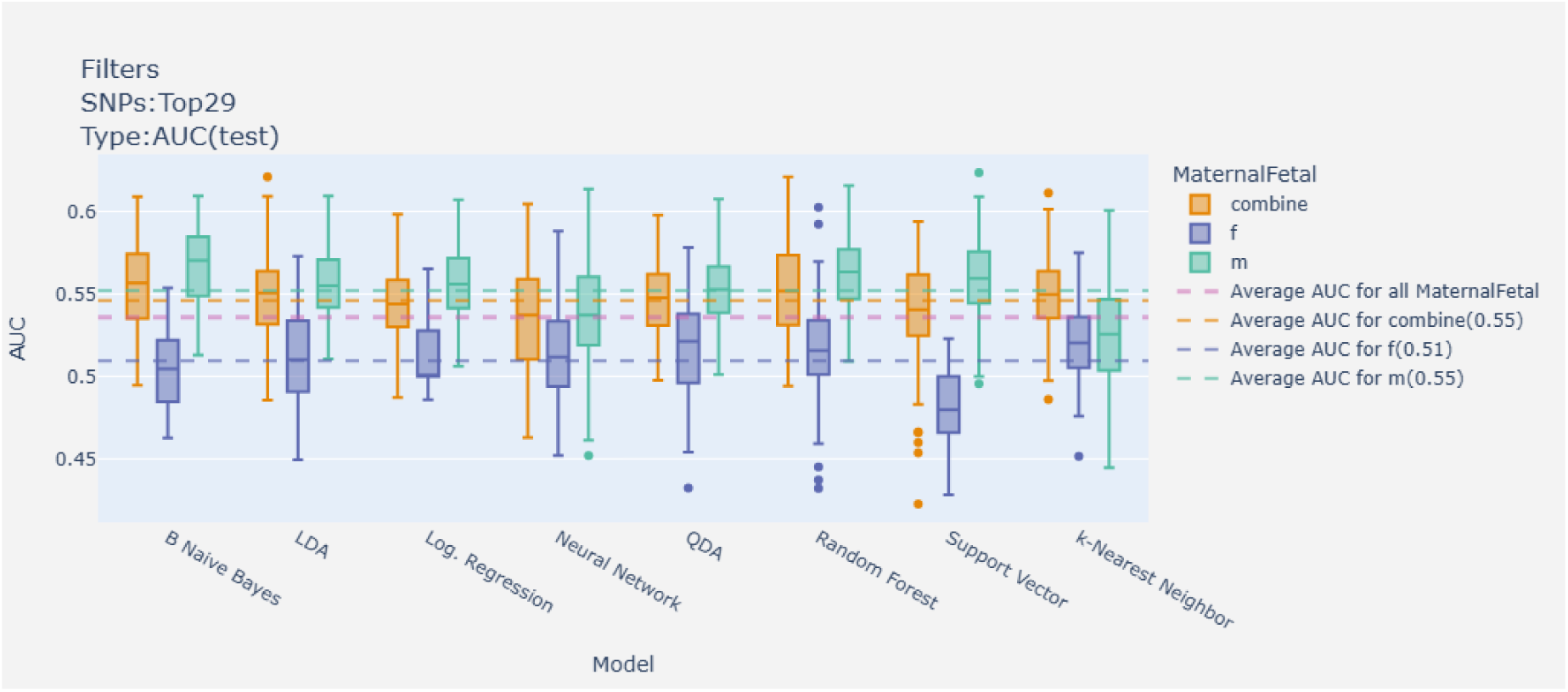
AUC for the fetal, maternal, and combined top 29 SNPs by model, respectively.

### Including SNPs beyond the top 29 slightly improve predictability for the fetal genome

While the fetal genome does not provide as much predictive power as the maternal genome for the top 29 SNPs, incorporating additional SNPs beyond the top ones does lead to a slight improvement in predictability, as seen in Fig. 6. This suggests that there are informative SNPs outside of the most recognized top SNPs that contribute to the fetal genome’s predictive ability. However, the overall results (AUCs around 0.54), are still lower than for the top 29 on the maternal genome.

**Figure 6:**
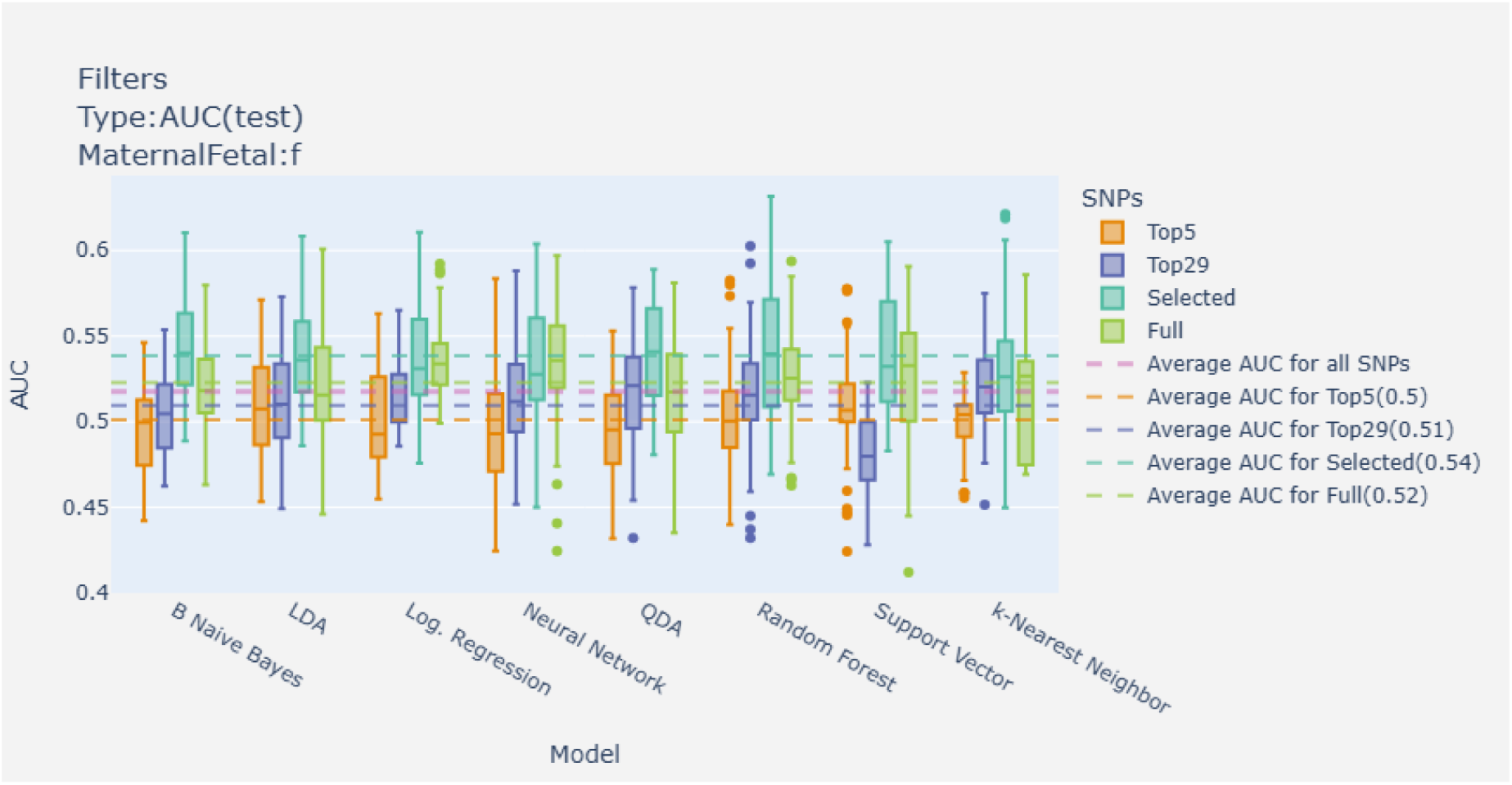
AUC by model for different sets of SNPs in the fetal genome.

## 4 Discussion

In this project, we aimed to investigate the capacity of machine learning models to predict PTB. The results indicated that more complex models or sets of SNPs did not significantly improve predictive accuracy, suggesting that non-additive effects of the identified loci may not be substantial enough to enhance predictions. This aligns with the results from a previous study, where non-additive effects showed no significant contribution to the phenotypic variance of 70 different traits [28].

Interestingly, expanding the analysis to include additional SNPs within the associated loci did not enhance predictive performance. This finding suggests that the lead SNPs hold the majority of the predictive power and that the inclusion of more SNPs may introduce noise rather than meaningful information, despite experimental studies suggesting that nearly all variants have a functional impact [29]. As has been previously shown for height [30], our results also highlight that gene-gene interactions are likely limited within known loci. It is noteworthy that limiting the analysis to the top five SNPs resulted in even poorer performance, reinforcing the notion that a broader selection of informative SNPs is necessary for effective predictive modeling. Hence, identifying novel SNPs associated with gestational duration or PTB might improve the prediction of PTB.

When considering the fetal genome, the results indicated a significantly lower predictive capacity compared to the maternal one. The combination of maternal and fetal genomes did slightly improve performance but did not yield better results than the maternal genome alone. This suggests that the loci identified through maternal genome studies are not as influential in the fetal genome context, despite some of these loci having either maternal and fetal effects or fetal effects only [13].

Preterm delivery is a multi-factorial condition, with heredity estimated to account for about 20-30% [9]–[12]. The genetic influence on preterm delivery may also differ between individuals and lack of clarity regarding which women in the cohort delivered preterm due to genetic factors further complicates the interpretation of the results. Including both environmental and genetic factors in the analysis could provide new essential information about the underlying mechanisms.

Our results highly disagree with a prior study,[14] addressed in the introduction, on predicting preterm delivery with machine learning models using genetic data. The study did not provide any code or information about the genetic variants used, hence we were not able to validate or compare results appropriately. We also acknowledge that the studies are conducted on different populations. However, given that gestational duration has a heritability of 20-30% [9]–[12], it is surprising that the previous study was able to predict preterm delivery with over 99% certainty based on maternal genetics alone.

The main limitation of our study is the inherent distribution of gestational duration and preterm delivery. The distribution between case and control samples is highly skewed in favor of controls, which can affect machine learning classifiers negatively. We have used random undersampling when training the models to avoid giving control samples too much weight. We have also employed rigorous iterations of the random undersampling, to mitigate the risk of losing valuable information in the process [31]. Using a continuous outcome could provide additional information, however, here we decided to focus on a clinically-relevant phenotype that is commonly used for decision-making.

For computational reasons, we limited the study to the gestational duration-associated regions identified in a previous study [13]. It is possible that including additional regions would have provided new information. However, including the whole genome is not feasible, and selecting the regions identified in the largest GWAS to date allowed us to answer whether there is additional predictive information beyond the top SNPs.

Since only genetic data was used in this study, we were unable to stratify between different subgroups of women delivering preterm. Different genetic variants may be more or less relevant for different subgroups. For instance, we have recently shown that the maternal genome has a larger effect on gestational duration in the first pregnancy compared to subsequent ones [32]. A previous study also showed that parity interacts with different risk factors and has a large effect on gestational duration variance [33]. In addition to the effects of parity, infecttion or inflammation status of the mother has also been shown to influence the risk of preterm birth [34]. These examples all demonstrate that using a more holistic approach may enhance the predictive accuracy of models for PTB and provide a more comprehensive understanding of its underlying causes.

## 5 Conclusion

The associated genetic variants explored in this study only show minor effects individually, no complex interactions, and poor predictive capacity.

Future research could explore different ways of selecting variants, such as functional relevance, in addition to expanding on the number of loci to include. Subsequently, incorporating other factors such as environmental exposures, lifestyle factors, or epigenetic modifications may also enhance predictability.

## Data Availability

The consent given by the participants does not permit public access of data at the individual level. Researchers can apply for access to data through helsedata.no. Access requires approval from the Regional Committees for Medical and Health Research Ethics in Norway and an agreement with MoBa.
Code developed for the analyses is available in a public repository (https://github.com/PerinatalLab/ml_genetics/) and upon request.

## Acknowledgements

We thank the Norwegian Institute of Public Health (NIPH) for generating high-quality genomic data. This research is part of the HARVEST collaboration, supported by the Research Council of Norway (#229624). We also thank the NORMENT Centre for providing genotype data, funded by the Research Council of Norway (#223273), South East Norway Versjon 6.9 3 Health Authorities and Stiftelsen Kristian Gerhard Jebsen. We further thank the Center for Diabetes Research, the University of Bergen for providing genotype data and performing quality control and imputation of the data funded by the ERC AdG project SELECTionPREDISPOSED, Stiftelsen Kristian Gerhard Jebsen, Trond Mohn Foundation, the Research Council of Norway, the Novo Nordisk Foundation, the University of Bergen, and the Western Norway Health Authorities. We are grateful to all the participating families in Norway who take part in this ongoing cohort study.

We are grateful for the funding from Lilla Barnets-foundation, awarded to Julius Juodakis, which helped support the project. The research activities were also made possible by funding received by Bo Jacobsson from The Swedish Research Council, Stockholm, Sweden (2019-01004 and 2024-02502), The Research Council of Norway, Oslo, Norway (FRIMEDBIO # 547711), March of Dimes (# 21-FY16-121), and the Agreement concerning research and education of doctors (ALFGBG-965353 and ALFGBG-1005151). As well as funding received by Pol Solé-Navais from The Swedish Research Council, Stockholm, Sweden (2023-02735), The Swedish Society for Medical Research (SG-24-0105-B) and the Agreement concerning research and education of doctors (ALFGBG-1005149).

## Acronyms

AUC: Area Under the Curve
B. Naive Bayes: Bernoulli Naive Bayes
FPR: False Positive Rate
GWAS: Genome-wide association study
IVF: In Vitro Fertilisation
LDA: Linear Discriminant Analysis
MBRN: Medical Birth Registry of Norway
MCC: Matthews Correlation Coefficient
MoBa: Norwegian Mother, Father and Child Cohort Study
OR: Odds Ratio
PRS: Polygenic Risk Score
PTB: Preterm Birth
QDA: Quadratic Discriminant Analysis
ROC: Receiver Operating Characteristic
SNP: Single Nucleotide Polymorphism
TPE: Tree-structured Parzen Estimator
TPR: True Positive Rate

## A Resampling Methods

**Figure S1:**
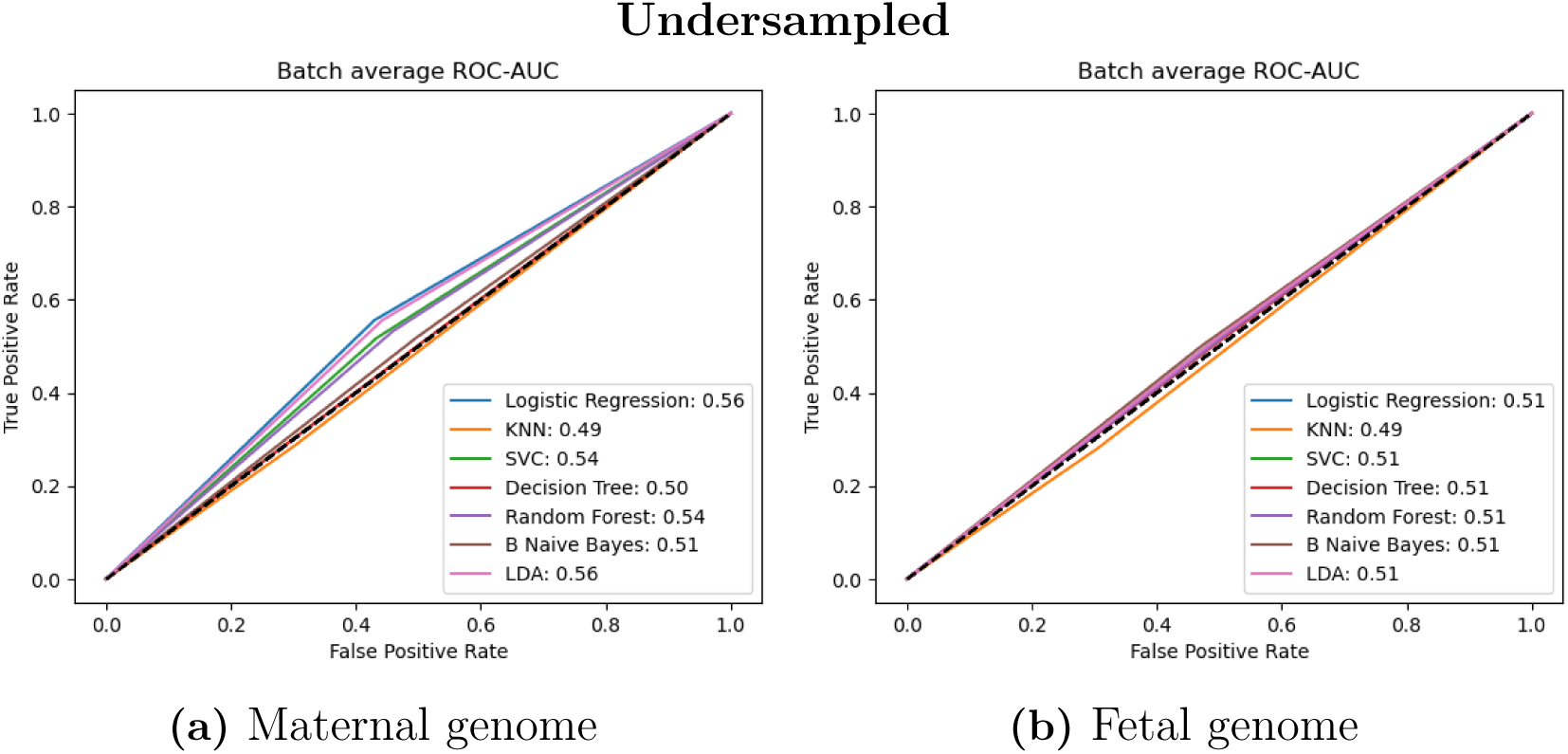
AUC for different models trained and tested on the top SNPs, with basic undersampling of training data

**Figure S2:**
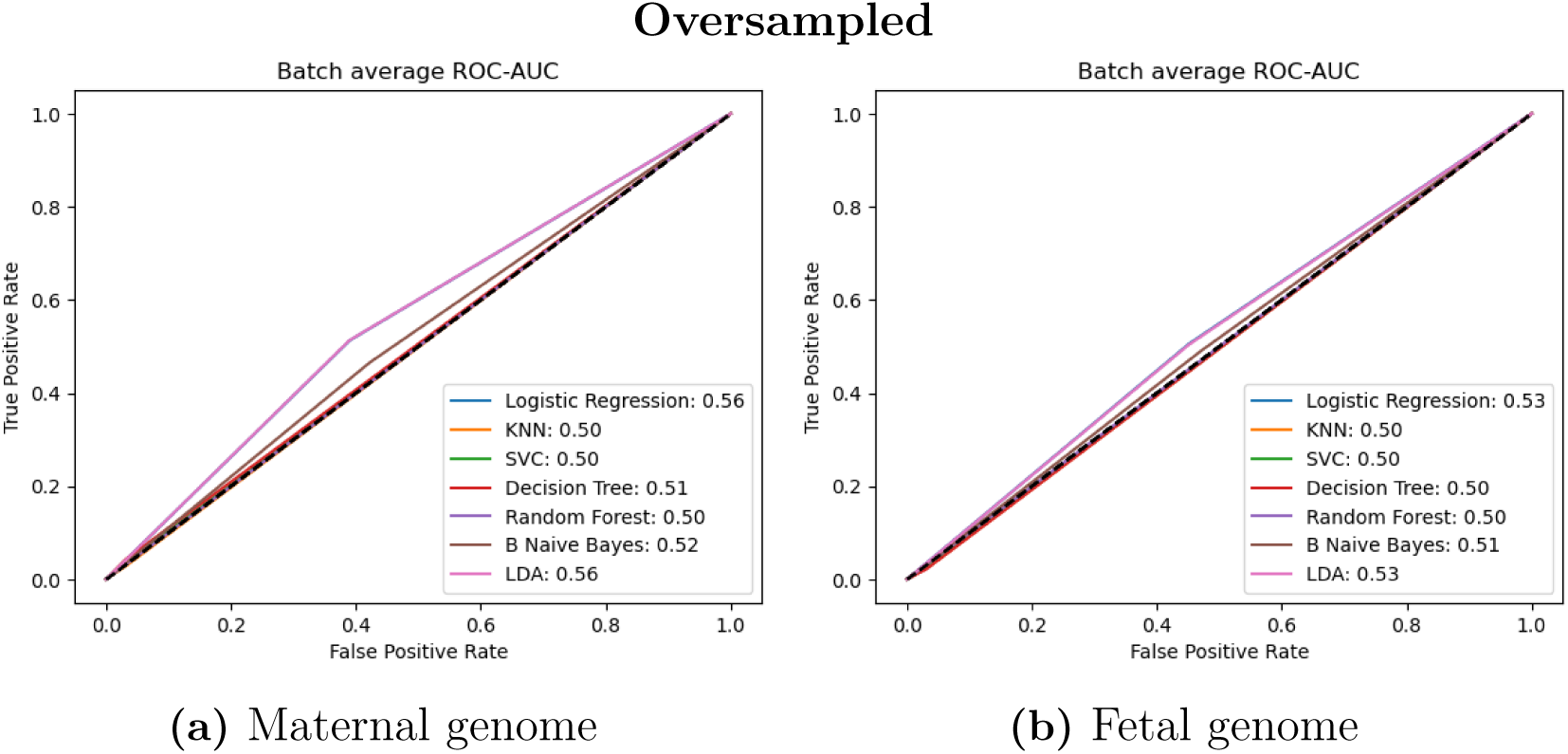
AUC for different models trained and tested on the top SNPs, with basic oversampling of training data

**Figure S3:**
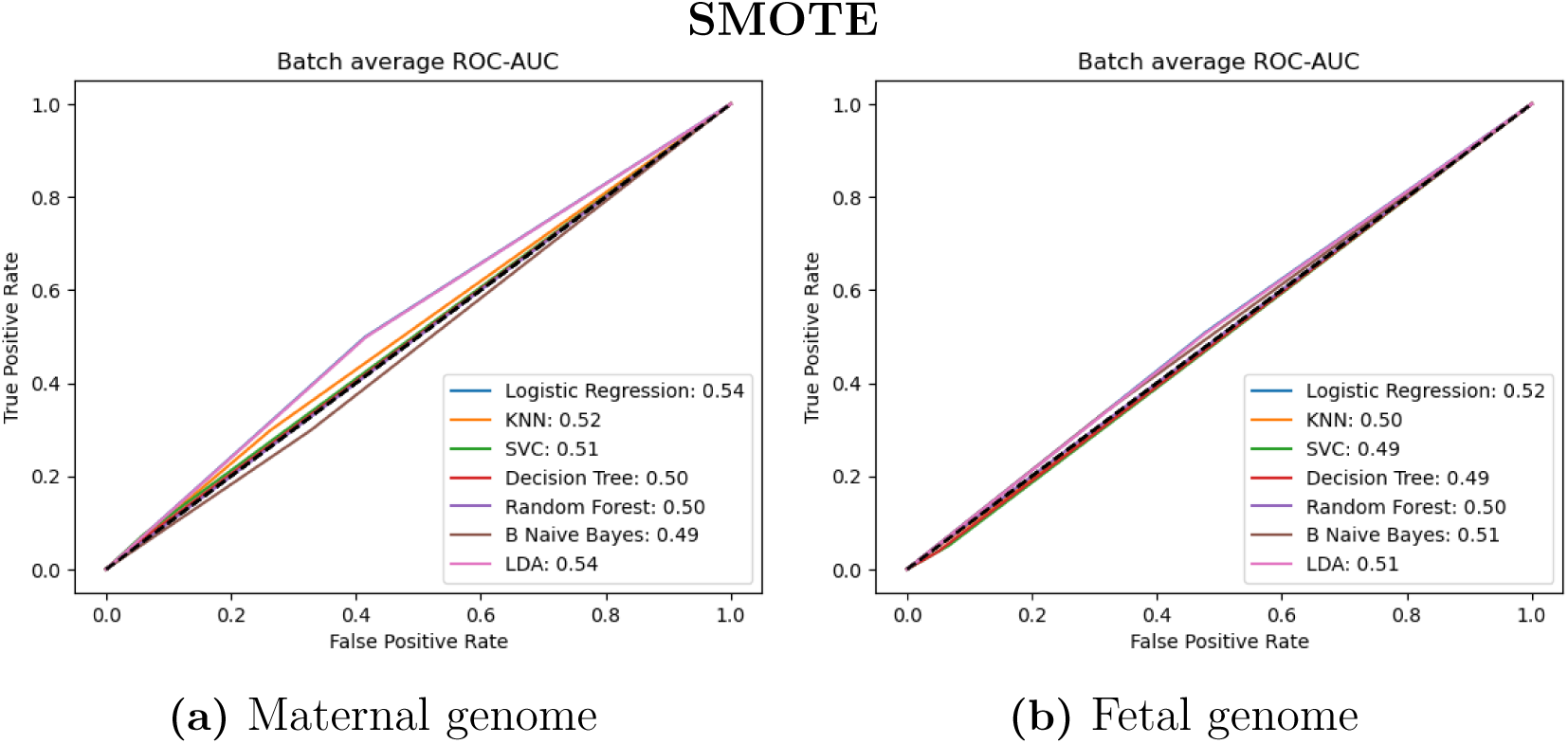
AUC for different models trained and tested on the top SNPs, with SMOTE oversampling of training data

**Figure S4:**
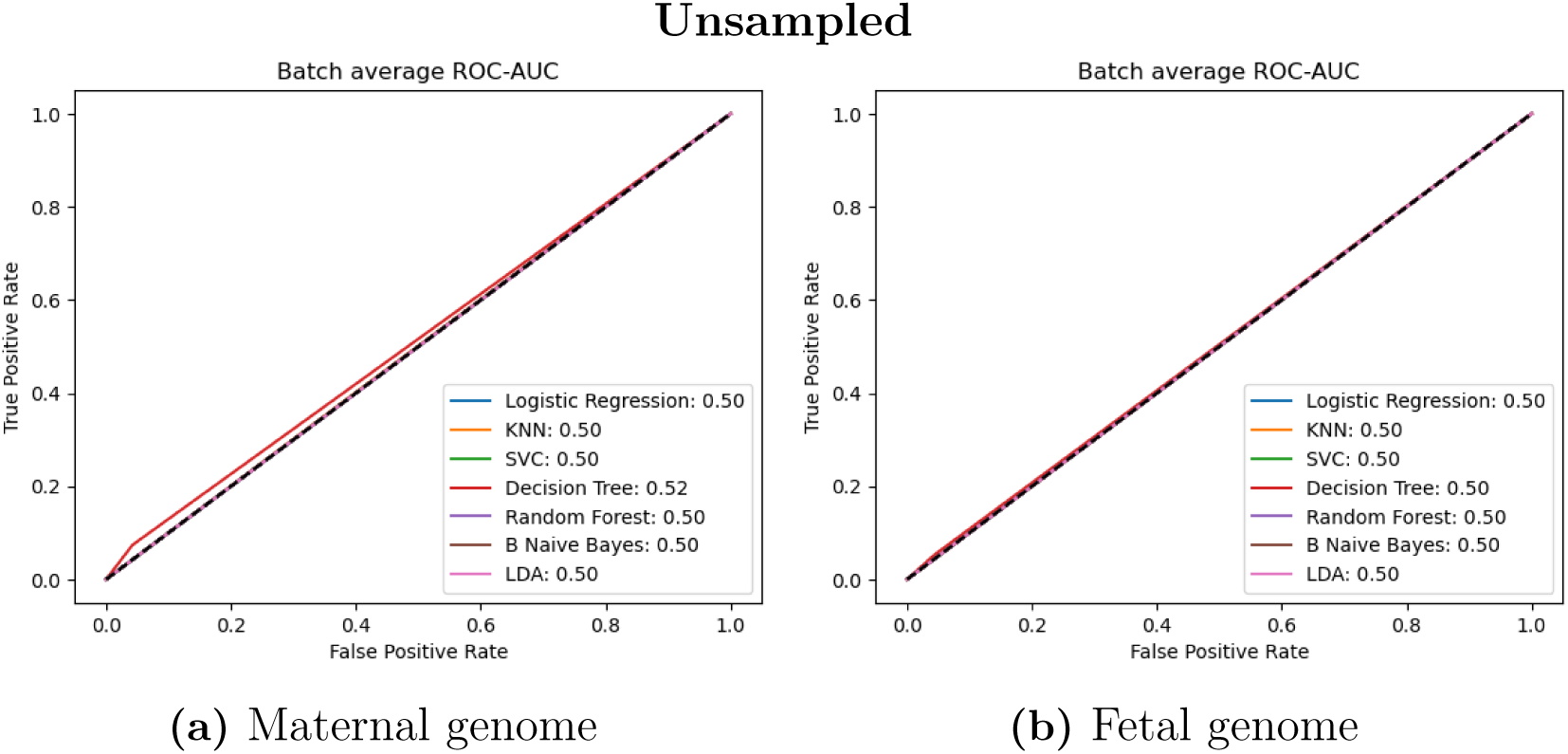
AUC for different models trained and tested on the top SNPs, without resampling of training data

## B Equations

### F-ANOVA

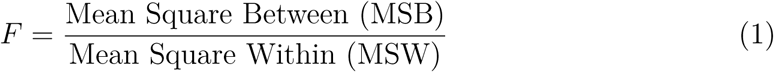

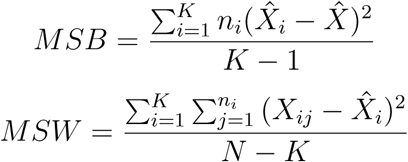

Where,

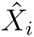 = sample mean of the ith group

*n_i_* = sample size of the ith group

*K* = number of groups

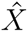= overall mean

*X_ij_* = jth observation in the ith group

*N* = Total number of observations/total sample size

### Area Under the ROC curve

Receiver Operating Characteristic (ROC) refers to the combination of Recall or True Positive Rate (TPR) and False Positive Rate (FPR) at different thresholds, *θ*. The two metrics are plotted against each other with the FPR on the x-axis and the TPR on the y-axis. The Area Under the ROC-curve, or AUC, summarizes the information in the ROC plot to a numeric value. “The AUC of a predictor f is defined as:” [35]

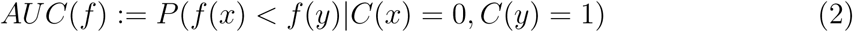

With the unbiased estimator:

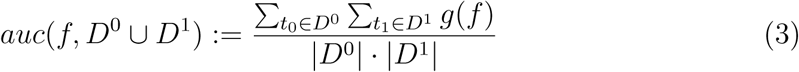

Where:

*D*^0^ is a set of negative examples

*D*^1^ is a set of positive examples

and

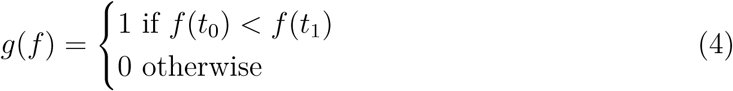

AUC include performance across all possible classification thresholds. It measures the ability of the classifier to distinguish between the positive and negative classes. The score can range between 0–1. however, a score of 0.5 suggests that the classifier can not distinguish one category from the other better than a random guess.

### Matthews Correlation Coefficient

The coefficient ranges from -1 to +1, where +1 is a perfect prediction, 0 is an average random prediction and -1 is a perfect inverse prediction.

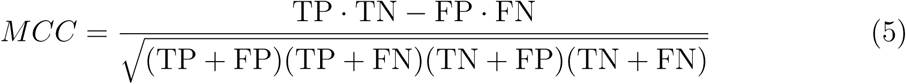

Where:

- TP (True Positive) is the number of true positive predictions.
- TN (True Negative) is the number of true negative predictions.
- FP (False Positive) is the number of false positive predictions.
- FN (False Negative) is the number of false negative predictions.

### Odds Ratio

The odds ratio (OR) measures the relative odds of an outcome, PTB, given a certain exposure, in this case, being classified as high risk by the model. An OR greater than 1 indicates a higher odds of the outcome occurring in the exposed group compared to the non-exposed group, while an OR less than 1 indicates a lower odds. An OR of exactly 1 suggests no association between the exposure and the outcome.

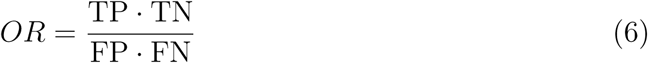

Where:

- TP (True Positive) is the number of true positive predictions.
- TN (True Negative) is the number of true negative predictions.
- FP (False Positive) is the number of false positive predictions.
- FN (False Negative) is the number of false negative predictions.

## C Supplementary tables

**Table S1:** Metrics by model on the Top29 subset in the fetal genome.

| Model | AUC |  |  | OR |  | Sens | Spec | MCC |
| --- | --- | --- | --- | --- | --- | --- | --- | --- |
|  | mean | median | 95% CI | mean | 95% CI | mean | mean | mean |
| B Naive Bayes | 0.503 | 0.505 | (0.5, 0.51) | 1.037 | (1.0, 1.07) | 0.501 | 0.505 | 0.002 |
| LDA | 0.512 | 0.510 | (0.51, 0.52) | 1.062 | (1.02, 1.1) | 0.504 | 0.507 | 0.003 |
| Log. Regression | 0.513 | 0.500 | (0.51, 0.52) | 0.608 | (0.49, 0.73) | 0.279 | 0.735 | 0.005 |
| Neural Network | 0.513 | 0.512 | (0.51, 0.52) | 1.100 | (1.05, 1.15) | 0.499 | 0.519 | 0.006 |
| QDA | 0.517 | 0.521 | (0.51, 0.52) | 1.114 | (1.07, 1.16) | 0.501 | 0.520 | 0.007 |
| Random Forest | 0.515 | 0.516 | (0.51, 0.52) | 1.132 | (1.09, 1.18) | 0.526 | 0.499 | 0.008 |
| Support Vector | 0.481 | 0.480 | (0.48, 0.49) | 1.123 | (1.08, 1.17) | 0.523 | 0.500 | 0.007 |
| k-Nearest Neighbor | 0.521 | 0.520 | (0.52, 0.53) | 1.212 | (1.14, 1.28) | 0.292 | 0.740 | 0.012 |

**Table S2:**
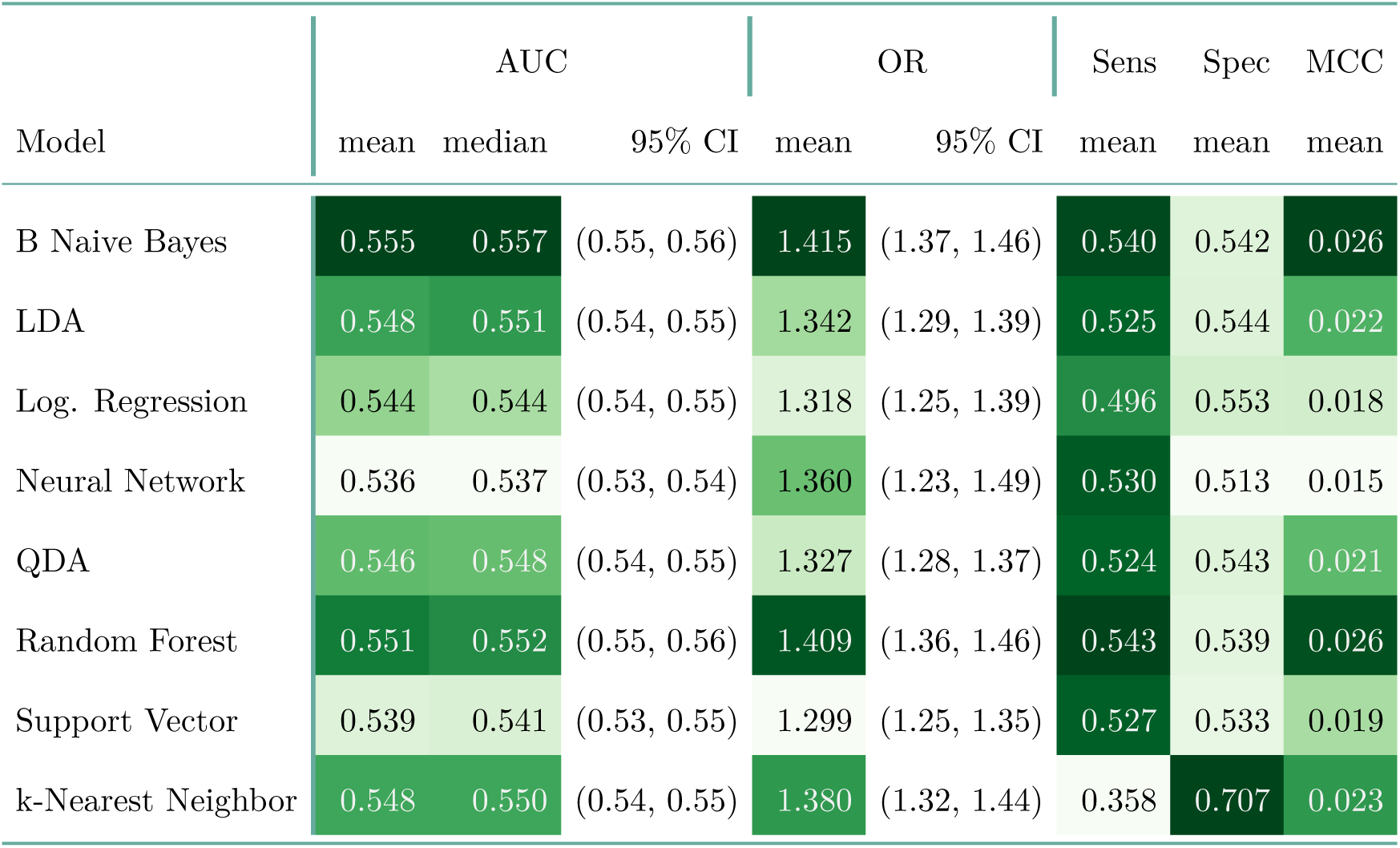
Metrics by model on the Top29 subset in the combined maternal and fetal genome.

**Table S3:** Metrics by model for the Selected subset in the maternal genome.

| Model | AUC |  |  | OR |  | Sens | Spec | MCC |
| --- | --- | --- | --- | --- | --- | --- | --- | --- |
|  | mean | median | 95% CI | mean | 95% CI | mean | mean | mean |
| B Naive Bayes | 0.545 | 0.537 | (0.54, 0.55) | 1.306 | (1.24, 1.38) | 0.335 | 0.717 | 0.018 |
| LDA | 0.533 | 0.533 | (0.53, 0.54) | 1.187 | (1.13, 1.24) | 0.313 | 0.720 | 0.011 |
| Log. Regression | 0.537 | 0.533 | (0.53, 0.54) | 1.211 | (1.15, 1.27) | 0.359 | 0.680 | 0.013 |
| Neural Network | 0.533 | 0.530 | (0.53, 0.54) | 1.217 | (1.14, 1.29) | 0.368 | 0.660 | 0.011 |
| QDA | 0.527 | 0.527 | (0.52, 0.53) | 1.355 | (1.29, 1.42) | 0.259 | 0.788 | 0.019 |
| Random Forest | 0.530 | 0.525 | (0.52, 0.54) | 1.205 | (1.15, 1.26) | 0.388 | 0.650 | 0.013 |
| Support Vector | 0.530 | 0.526 | (0.52, 0.54) | 1.170 | (1.11, 1.23) | 0.315 | 0.714 | 0.010 |
| k-Nearest Neighbor | 0.525 | 0.535 | (0.52, 0.53) | 1.303 | (1.12, 1.49) | 0.043 | 0.964 | 0.007 |

**Table S4:** Metrics by model on the Selected subset in the fetal genome.

| Model | AUC |  |  | OR |  | Sens | Spec | MCC |
| --- | --- | --- | --- | --- | --- | --- | --- | --- |
|  | mean | median | 95% CI | mean | 95% CI | mean | mean | mean |
| B Naive Bayes | 0.544 | 0.540 | (0.54, 0.55) | 1.361 | (1.32, 1.41) | 0.346 | 0.718 | 0.023 |
| LDA | 0.541 | 0.536 | (0.53, 0.55) | 1.347 | (1.3, 1.4) | 0.348 | 0.714 | 0.022 |
| Log. Regression | 0.537 | 0.531 | (0.53, 0.54) | 1.279 | (1.23, 1.33) | 0.372 | 0.681 | 0.018 |
| Neural Network | 0.536 | 0.528 | (0.53, 0.54) | 1.252 | (1.18, 1.32) | 0.383 | 0.663 | 0.016 |
| QDA | 0.540 | 0.541 | (0.53, 0.55) | 1.360 | (1.3, 1.42) | 0.274 | 0.780 | 0.021 |
| Random Forest | 0.541 | 0.540 | (0.53, 0.55) | 1.366 | (1.29, 1.44) | 0.401 | 0.665 | 0.022 |
| Support Vector | 0.540 | 0.532 | (0.53, 0.55) | 1.322 | (1.27, 1.38) | 0.348 | 0.709 | 0.020 |
| k-Nearest Neighbor | 0.529 | 0.526 | (0.52, 0.54) | 1.322 | (1.17, 1.47) | 0.076 | 0.934 | 0.008 |

**Table S5:**
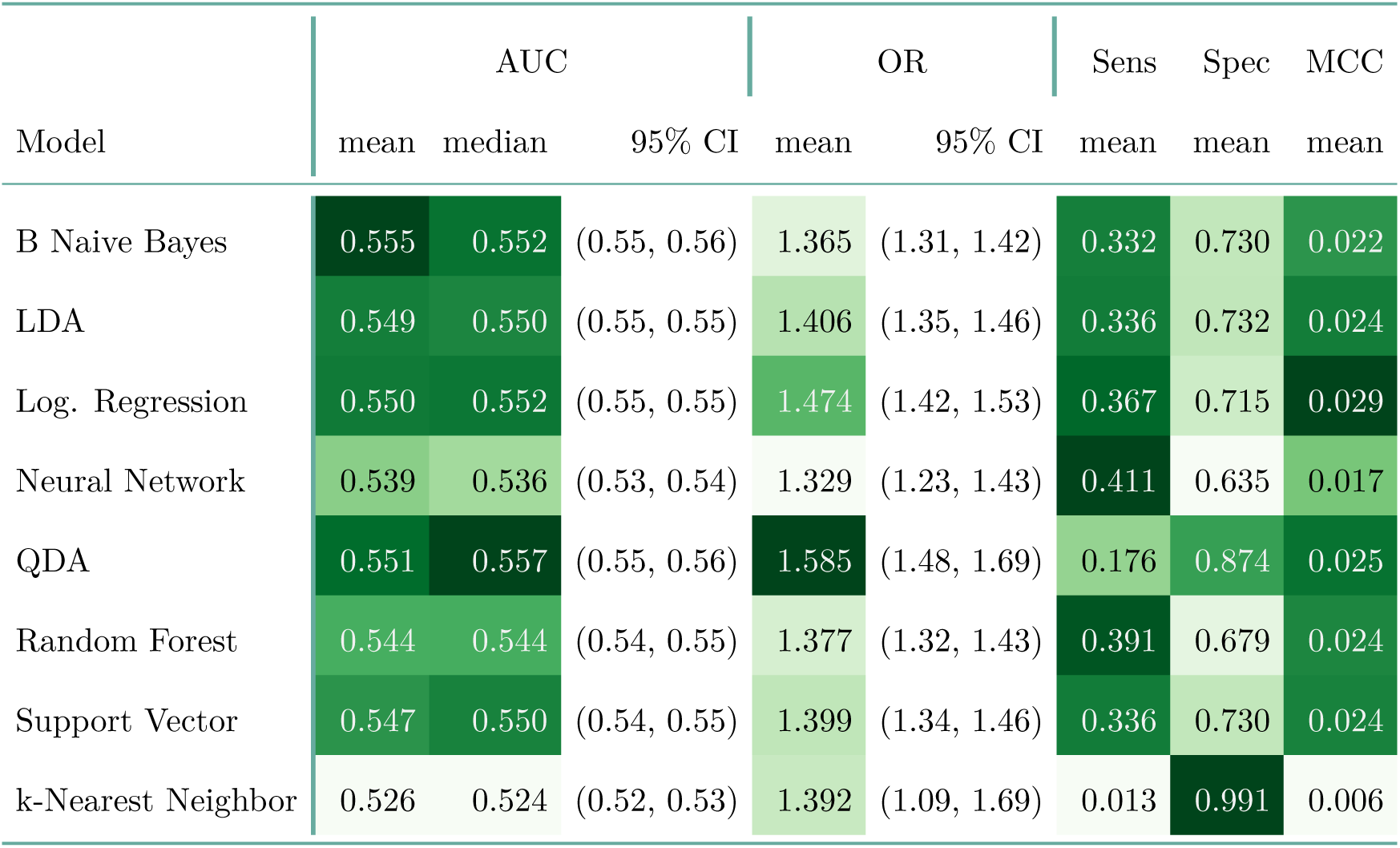
Metrics by model on the Selected subset in the combined maternal and fetal genome.

| Model | AUC |  |  | OR |  | Sens | Spec | MCC |
| --- | --- | --- | --- | --- | --- | --- | --- | --- |
|  | mean | median | 95% CI | mean | 95% CI | mean | mean | mean |
| B Naive Bayes | 0.555 | 0.552 | (0.55, 0.56) | 1.365 | (1.31, 1.42) | 0.332 | 0.730 | 0.022 |
| LDA | 0.549 | 0.550 | (0.55, 0.55) | 1.406 | (1.35, 1.46) | 0.336 | 0.732 | 0.024 |
| Log. Regression | 0.550 | 0.552 | (0.55, 0.55) | 1.474 | (1.42, 1.53) | 0.367 | 0.715 | 0.029 |
| Neural Network | 0.539 | 0.536 | (0.53, 0.54) | 1.329 | (1.23, 1.43) | 0.411 | 0.635 | 0.017 |
| QDA | 0.551 | 0.557 | (0.55, 0.56) | 1.585 | (1.48, 1.69) | 0.176 | 0.874 | 0.025 |
| Random Forest | 0.544 | 0.544 | (0.54, 0.55) | 1.377 | (1.32, 1.43) | 0.391 | 0.679 | 0.024 |
| Support Vector | 0.547 | 0.550 | (0.54, 0.55) | 1.399 | (1.34, 1.46) | 0.336 | 0.730 | 0.024 |
| k-Nearest Neighbor | 0.526 | 0.524 | (0.52, 0.53) | 1.392 | (1.09, 1.69) | 0.013 | 0.991 | 0.006 |

**Table S6:** Metrics by model in the Top5 subset on the maternal genome.

| Model | AUC |  |  | OR |  | Sens | Spec | MCC |
| --- | --- | --- | --- | --- | --- | --- | --- | --- |
|  | mean | median | 95% CI | mean | 95% CI | mean | mean | mean |
| B Naive Bayes | 0.555 | 0.553 | (0.55, 0.56) | 1.371 | (1.31, 1.43) | 0.481 | 0.590 | 0.023 |
| LDA | 0.559 | 0.556 | (0.55, 0.56) | 1.477 | (1.41, 1.54) | 0.491 | 0.598 | 0.029 |
| Log. Regression | 0.559 | 0.560 | (0.55, 0.56) | 1.447 | (1.38, 1.51) | 0.484 | 0.599 | 0.027 |
| Neural Network | 0.514 | 0.508 | (0.51, 0.52) | 0.872 | (0.74, 1.01) | 0.456 | 0.571 | 0.010 |
| QDA | 0.546 | 0.546 | (0.54, 0.55) | 1.415 | (1.34, 1.49) | 0.402 | 0.670 | 0.025 |
| Random Forest | 0.545 | 0.540 | (0.54, 0.55) | 1.321 | (1.26, 1.38) | 0.480 | 0.582 | 0.020 |
| Support Vector | 0.535 | 0.537 | (0.53, 0.54) | 1.193 | (1.13, 1.26) | 0.552 | 0.483 | 0.012 |
| k-Nearest Neighbor | 0.531 | 0.527 | (0.52, 0.54) | 1.336 | (1.25, 1.42) | 0.141 | 0.886 | 0.014 |

**Table S7:** Metrics by model on the Top5 subset in the fetal genome.

| Model | AUC |  |  | OR |  | Sens | Spec | MCC |
| --- | --- | --- | --- | --- | --- | --- | --- | --- |
|  | mean | median | 95% CI | mean | 95% CI | mean | mean | mean |
| B Naive Bayes | 0.495 | 0.500 | (0.49, 0.5) | 0.983 | (0.94, 1.03) | 0.433 | 0.561 | -0.002 |
| LDA | 0.509 | 0.507 | (0.5, 0.51) | 1.080 | (1.04, 1.12) | 0.470 | 0.545 | 0.005 |
| Log. Regression | 0.502 | 0.493 | (0.5, 0.51) | 1.015 | (0.97, 1.06) | 0.447 | 0.556 | 0.001 |
| Neural Network | 0.495 | 0.493 | (0.49, 0.5) | 0.318 | (0.22, 0.41) | 0.630 | 0.361 | -0.003 |
| QDA | 0.495 | 0.495 | (0.49, 0.5) | 1.005 | (0.97, 1.04) | 0.388 | 0.610 | -0.001 |
| Random Forest | 0.503 | 0.500 | (0.5, 0.51) | 1.019 | (0.98, 1.06) | 0.510 | 0.491 | 0.000 |
| Support Vector | 0.512 | 0.507 | (0.51, 0.52) | 0.721 | (0.61, 0.83) | 0.829 | 0.164 | -0.004 |
| k-Nearest Neighbor | 0.500 | 0.504 | (0.5, 0.5) | 0.783 | (0.67, 0.89) | 0.082 | 0.903 | -0.008 |

**Table S8:** Metrics by model on the Top5 subset in the combined maternal and fetal genome.

| Model | AUC |  |  | OR |  | Sens | Spec | MCC |
| --- | --- | --- | --- | --- | --- | --- | --- | --- |
|  | mean | median | 95% CI | mean | 95% CI | mean | mean | mean |
| B Naive Bayes | 0.550 | 0.548 | (0.54, 0.56) | 1.368 | (1.31, 1.43) | 0.494 | 0.576 | 0.023 |
| LDA | 0.551 | 0.554 | (0.55, 0.56) | 1.388 | (1.33, 1.44) | 0.502 | 0.574 | 0.024 |
| Log. Regression | 0.547 | 0.546 | (0.54, 0.55) | 1.383 | (1.29, 1.48) | 0.506 | 0.556 | 0.021 |
| Neural Network | 0.523 | 0.520 | (0.52, 0.53) | 1.164 | (1.11, 1.22) | 0.460 | 0.568 | 0.009 |
| QDA | 0.528 | 0.526 | (0.52, 0.53) | 1.209 | (1.15, 1.27) | 0.466 | 0.574 | 0.013 |
| Random Forest | 0.531 | 0.531 | (0.52, 0.54) | 1.200 | (1.15, 1.25) | 0.498 | 0.541 | 0.012 |
| Support Vector | 0.532 | 0.537 | (0.53, 0.54) | 0.989 | (0.88, 1.09) | 0.646 | 0.381 | 0.004 |
| k-Nearest Neighbor | 0.519 | 0.519 | (0.51, 0.53) | 0.919 | (0.84, 1.0) | 0.094 | 0.898 | -0.005 |

## D Supplementary figures

**Figure S5:**
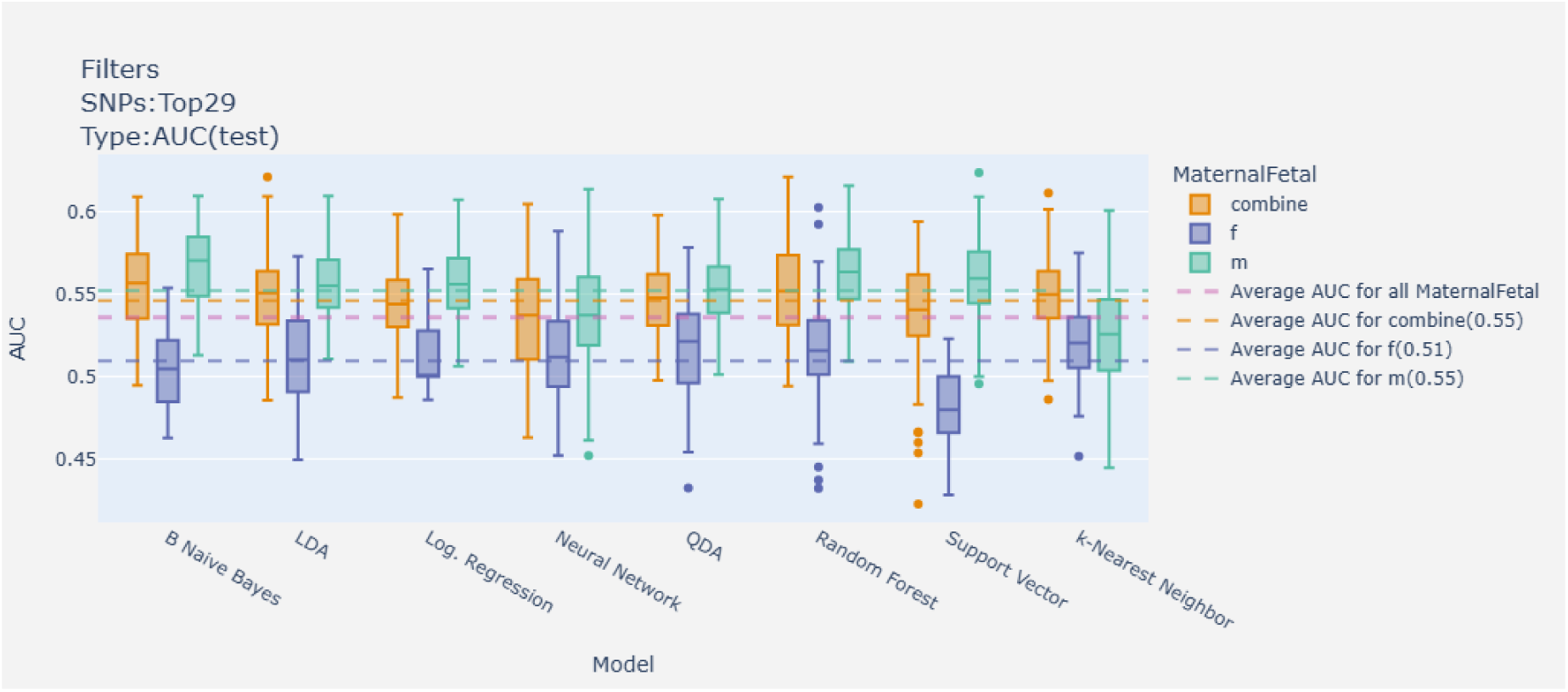
AUC for the fetal, maternal, and combined Top29 SNPs by model, respectively.

**Figure S6:**
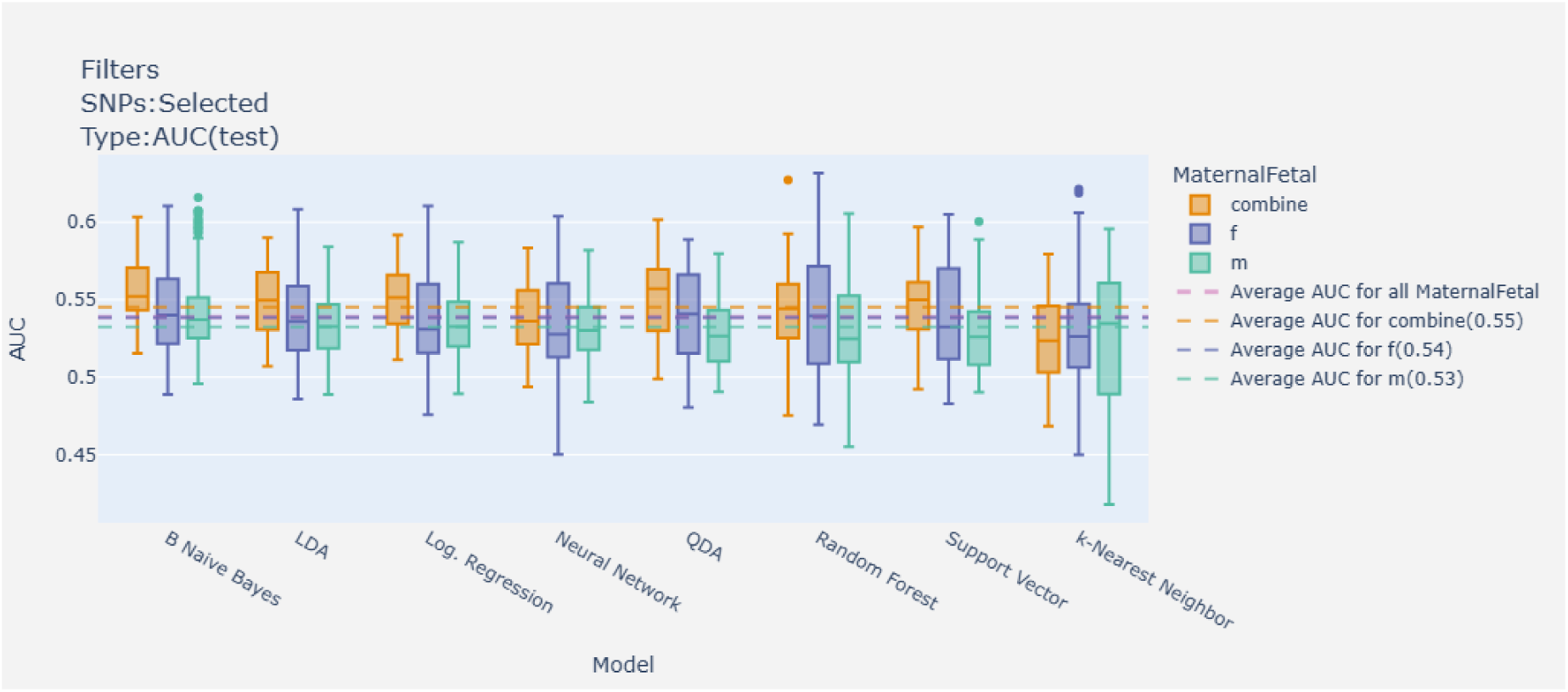
AUC for the fetal, maternal, and combined selected SNPs by model, respectively.

**Figure S7:**
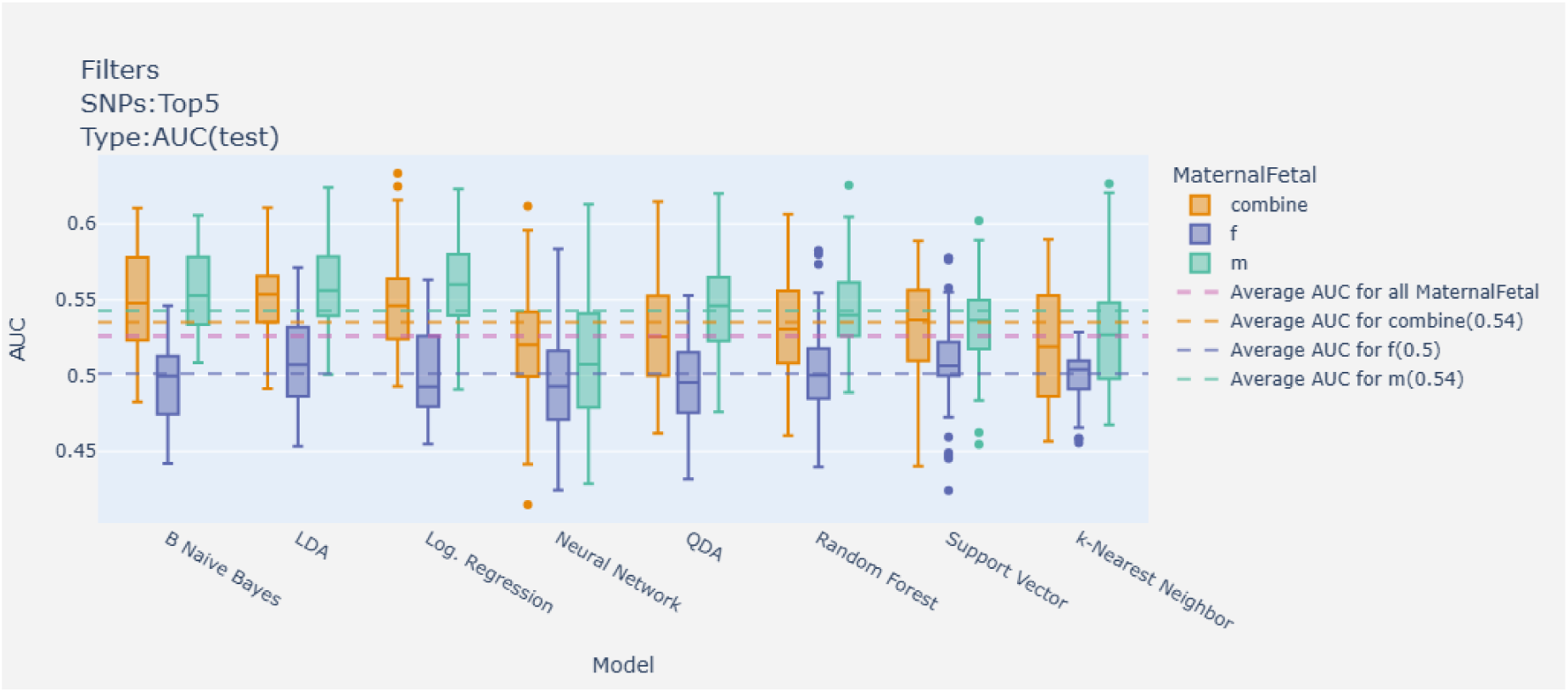
AUC for the fetal, maternal, and combined Top5 SNPs by model, respectively.

**Figure S8:**
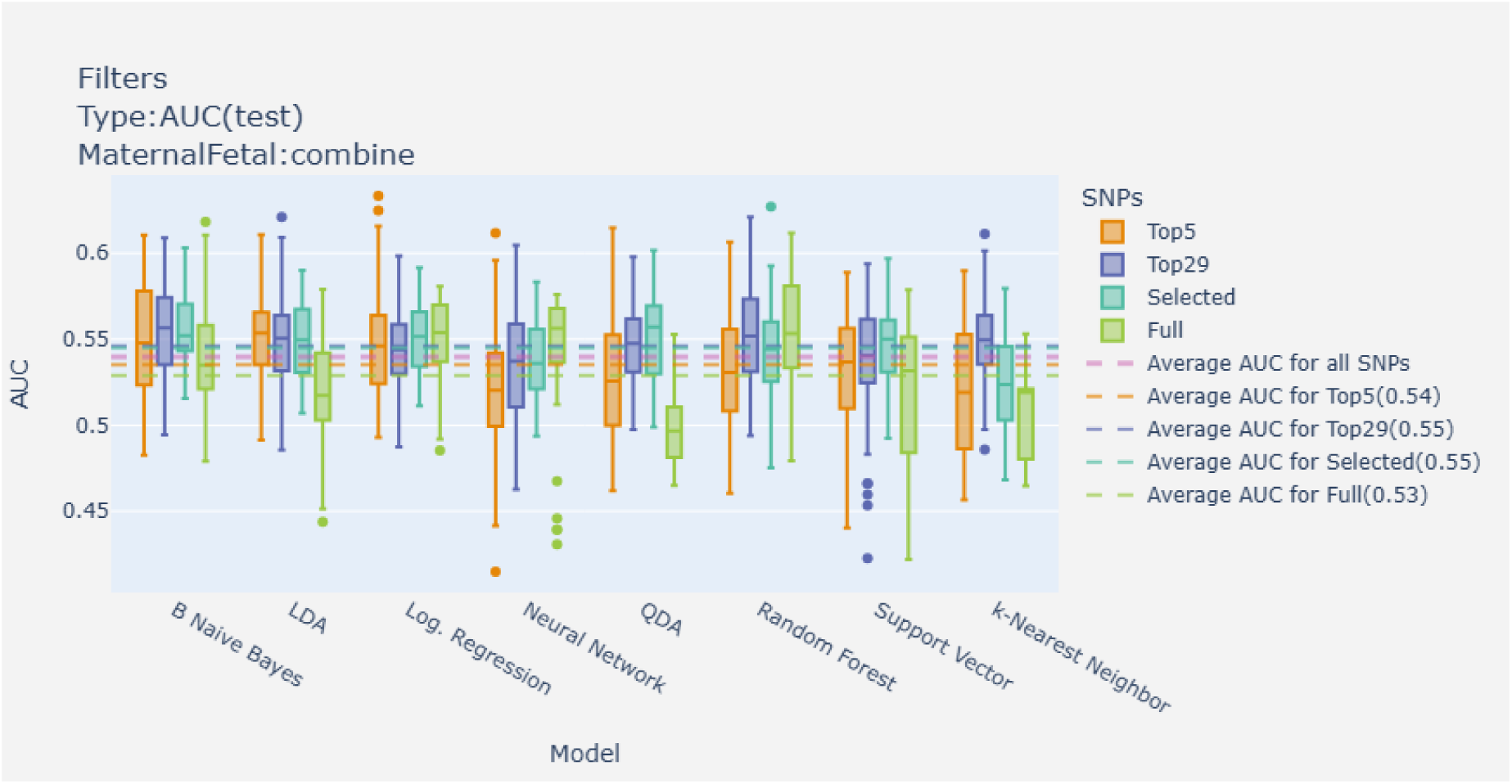
AUC by model on different sets of SNPs in the combined maternal and fetal genome.

## E Variants

**Table S9:**
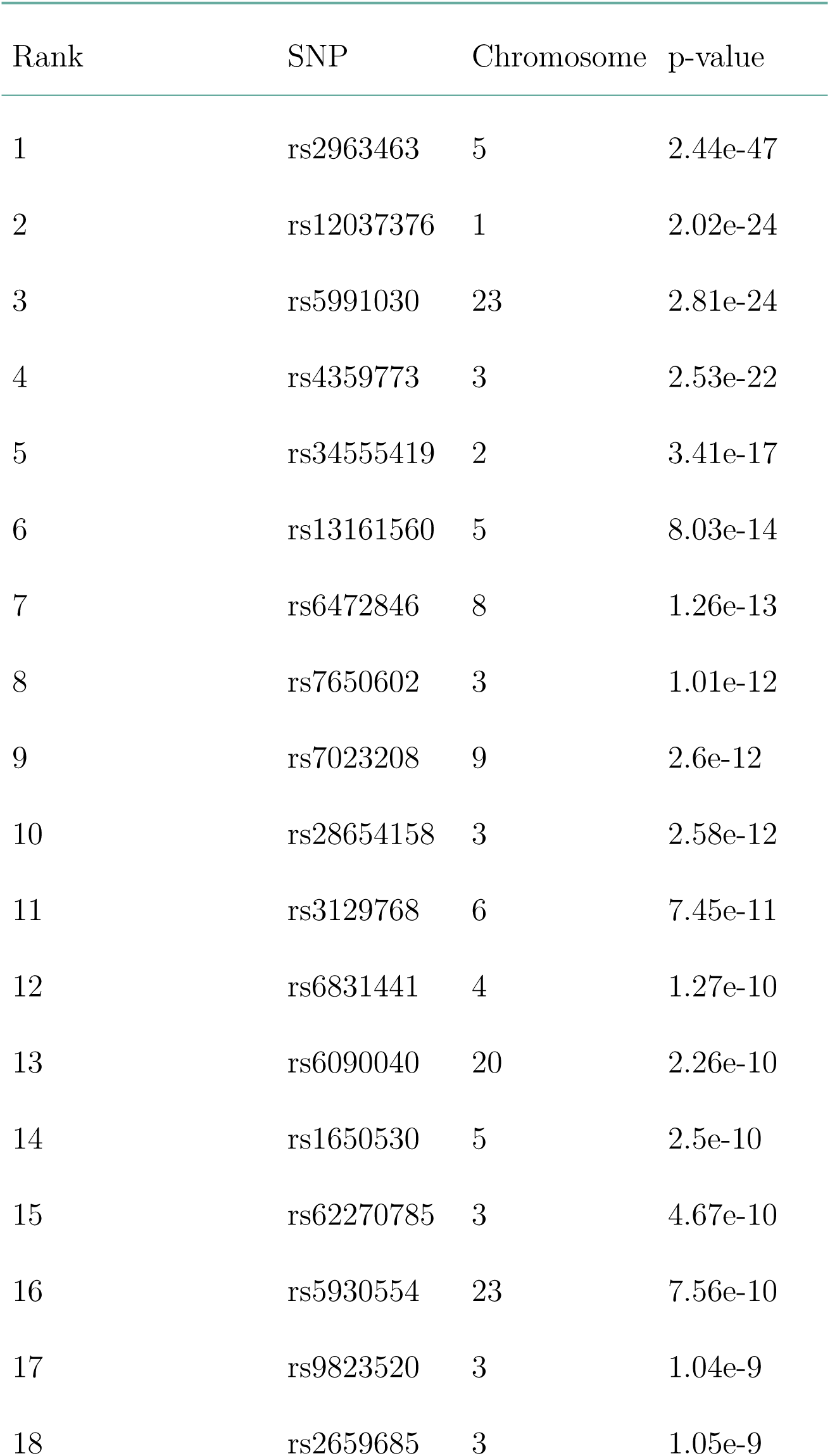
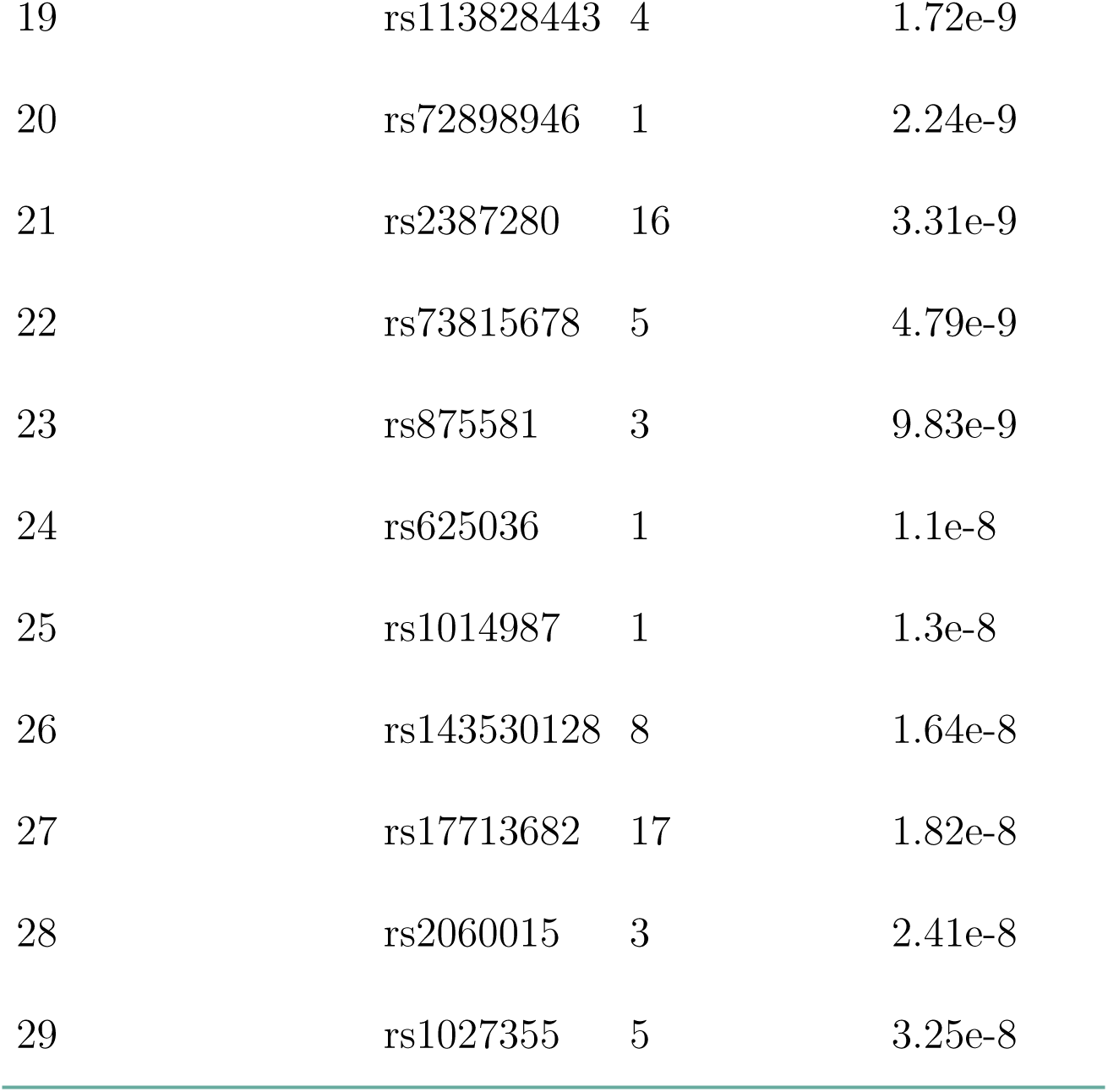
Top 29 SNPs in the gestational duration-associated loci, ranked by p-value.

**Table S10:** Selected SNPs, maternal genome. List of the SNPs in the Selected subset for the maternal genome, sorted by p-value. Rank is calculated over all folds and the total number of folds where the SNP is included is listed under Total.

| Rank | SNP | Chromosome | p-value | Fold |  |  |  |  | Total |
| --- | --- | --- | --- | --- | --- | --- | --- | --- | --- |
|  |  |  |  | 0 | 1 | 2 | 3 | 4 |  |
| 203 | rs2963463 | 5 | 2.44e-47 | 0 | 0 | 0 | 1 | 0 | 1 |
| 1 | rs34555419 | 2 | 3.41e-17 | 1 | 1 | 1 | 1 | 1 | 5 |
| 64 | rs7023208 | 9 | 2.60e-12 | 1 | 1 | 1 | 1 | 1 | 5 |

**Table S10:** Selected SNPs, maternal genome.
| Rank | SNP | Chromosome | p-value | Fold |  |  |  |  | Total |
| --- | --- | --- | --- | --- | --- | --- | --- | --- | --- |
|  |  |  |  | 0 | 1 | 2 | 3 | 4 |  |
| 149 | rs143530128 | 8 | 1.64e-08 | 1 | 0 | 0 | 0 | 1 | 2 |
| 34 | rs116449362 | 4 | 5.46e-07 | 1 | 1 | 1 | 0 | 0 | 3 |
| 92 | rs36096565 | 6 | 9.70e-07 | 0 | 1 | 1 | 0 | 0 | 2 |
| 13 | rs58218495 | 3 | 1.09e-06 | 1 | 1 | 0 | 1 | 1 | 4 |
| 29 | rs150703020 | 1 | 6.03e-06 | 0 | 0 | 1 | 1 | 0 | 2 |
| 188 | rs34776314 | 4 | 1.12e-05 | 0 | 0 | 0 | 1 | 0 | 1 |
| 191 | rs7694130 | 4 | 1.45e-05 | 0 | 0 | 1 | 0 | 0 | 1 |
| 194 | rs75506800 | 9 | 1.53e-05 | 1 | 0 | 0 | 0 | 1 | 2 |
| 58 | rs9270599 | 6 | 1.87e-05 | 1 | 0 | 1 | 1 | 0 | 3 |
| 73 | rs880466 | 1 | 2.42e-05 | 0 | 0 | 0 | 0 | 1 | 1 |
| 201 | rs406150 | 5 | 4.00e-05 | 0 | 0 | 0 | 1 | 0 | 1 |
| 79 | rs10917196 | 1 | 4.87e-05 | 0 | 0 | 0 | 1 | 0 | 1 |
| 207 | rs6457509 | 6 | 1.32e-04 | 0 | 1 | 0 | 0 | 0 | 1 |
| 25 | rs11589432 | 1 | 3.86e-04 | 0 | 1 | 0 | 0 | 1 | 2 |
| 3 | rs664777 | 1 | 4.41e-04 | 1 | 1 | 1 | 1 | 0 | 4 |
| 95 | rs145459800 | 2 | 7.97e-04 | 0 | 1 | 0 | 0 | 0 | 1 |

**Table S10:** Selected SNPs, maternal genome.
| Rank | SNP | Chromosome | p-value | Fold |  |  |  |  | Total |
| --- | --- | --- | --- | --- | --- | --- | --- | --- | --- |
|  |  |  |  | 0 | 1 | 2 | 3 | 4 |  |
| 124 | rs11707570 | 3 | 1.08e-03 | 0 | 0 | 1 | 0 | 0 | 1 |
| 133 | rs9872517 | 3 | 1.51e-03 | 0 | 0 | 1 | 0 | 0 | 1 |
| 140 | rs9839825 | 3 | 1.58e-03 | 0 | 0 | 0 | 1 | 0 | 1 |
| 146 | rs295307 | 3 | 2.44e-03 | 0 | 0 | 0 | 1 | 0 | 1 |
| 56 | rs187432070 | 3 | 2.76e-03 | 1 | 0 | 1 | 0 | 0 | 2 |
| 114 | rs111582361 | 3 | 3.55e-03 | 0 | 0 | 0 | 0 | 1 | 1 |
| 125 | rs10755076 | 3 | 4.16e-03 | 0 | 0 | 0 | 1 | 0 | 1 |
| 244 | rs201188737 | 16 | 4.36e-03 | 1 | 0 | 0 | 0 | 0 | 1 |
| 193 | rs10087246 | 8 | 4.46e-03 | 0 | 1 | 0 | 0 | 1 | 2 |
| 172 | rs116562818 | 3 | 4.64e-03 | 0 | 0 | 0 | 0 | 1 | 1 |
| 291 | rs141594674 | 23 | 5.18e-03 | 0 | 0 | 1 | 0 | 0 | 1 |
| 205 | rs79978285 | 5 | 5.18e-03 | 0 | 0 | 0 | 1 | 0 | 1 |
| 156 | rs113896410 | 3 | 5.55e-03 | 0 | 1 | 0 | 0 | 0 | 1 |
| 152 | rs72762640 | 9 | 5.76e-03 | 1 | 0 | 1 | 0 | 1 | 3 |
| 62 | rs142288650 | 4 | 6.34e-03 | 0 | 1 | 0 | 1 | 0 | 2 |
| 10 | rs79220368 | 4 | 6.54e-03 | 1 | 1 | 1 | 1 | 1 | 5 |

**Table S10:** Selected SNPs, maternal genome.
| Rank | SNP | Chromosome | p-value | Fold |  |  |  |  | Total |
| --- | --- | --- | --- | --- | --- | --- | --- | --- | --- |
|  |  |  |  | 0 | 1 | 2 | 3 | 4 |  |
| 63 | rs115701023 | 4 | 7.47e-03 | 1 | 0 | 1 | 0 | 0 | 2 |
| 45 | rs12635744 | 3 | 8.92e-03 | 0 | 0 | 1 | 0 | 1 | 2 |
| 166 | rs77984205 | 3 | 9.60e-03 | 0 | 0 | 0 | 0 | 1 | 1 |
| 61 | rs1401910 | 4 | 1.07e-02 | 1 | 0 | 0 | 0 | 1 | 2 |
| 129 | rs614664 | 3 | 1.22e-02 | 0 | 0 | 0 | 1 | 0 | 1 |
| 147 | rs79318144 | 8 | 1.22e-02 | 0 | 1 | 0 | 0 | 1 | 2 |
| 183 | rs141051250 | 5 | 1.23e-02 | 1 | 0 | 0 | 0 | 0 | 1 |
| 298 | rs73222943 | 23 | 1.28e-02 | 0 | 0 | 0 | 1 | 0 | 1 |
| 18 | rs9854729 | 3 | 1.29e-02 | 0 | 1 | 1 | 0 | 1 | 3 |
| 42 | rs9917792 | 3 | 1.34e-02 | 0 | 1 | 1 | 0 | 0 | 2 |
| 59 | rs114706200 | 3 | 1.39e-02 | 0 | 1 | 1 | 0 | 0 | 2 |
| 150 | rs9302815 | 16 | 1.51e-02 | 1 | 0 | 0 | 1 | 1 | 3 |
| 228 | rs2499767 | 6 | 1.51e-02 | 0 | 0 | 0 | 1 | 0 | 1 |
| 16 | rs2844580 | 6 | 1.61e-02 | 1 | 1 | 1 | 1 | 1 | 5 |
| 251 | rs78336371 | 17 | 1.74e-02 | 1 | 0 | 1 | 0 | 1 | 3 |
| 295 | rs145863381 | 23 | 1.78e-02 | 0 | 1 | 0 | 0 | 0 | 1 |

**Table S10:** Selected SNPs, maternal genome.
| Rank | SNP | Chromosome | p-value | Fold |  |  |  |  | Total |
| --- | --- | --- | --- | --- | --- | --- | --- | --- | --- |
|  |  |  |  | 0 | 1 | 2 | 3 | 4 |  |
| 37 | rs118027082 | 8 | 1.81e-02 | 1 | 1 | 1 | 0 | 1 | 4 |
| 121 | rs9825582 | 3 | 2.01e-02 | 0 | 0 | 0 | 1 | 0 | 1 |
| 71 | rs150834019 | 8 | 2.03e-02 | 1 | 0 | 0 | 1 | 1 | 3 |
| 180 | rs35794567 | 4 | 2.04e-02 | 0 | 0 | 0 | 1 | 0 | 1 |
| 177 | rs112199365 | 4 | 2.14e-02 | 0 | 0 | 0 | 1 | 0 | 1 |
| 20 | rs78793891 | 3 | 2.17e-02 | 0 | 0 | 1 | 1 | 1 | 3 |
| 96 | rs4684995 | 3 | 2.17e-02 | 1 | 0 | 0 | 0 | 0 | 1 |
| 200 | rs2269476 | 6 | 2.21e-02 | 1 | 0 | 0 | 0 | 0 | 1 |
| 187 | rs17360144 | 4 | 2.22e-02 | 0 | 0 | 0 | 0 | 1 | 1 |
| 21 | rs116825850 | 1 | 2.32e-02 | 1 | 0 | 1 | 0 | 0 | 2 |
| 297 | rs189441177 | 23 | 2.39e-02 | 0 | 0 | 0 | 0 | 1 | 1 |
| 204 | rs559313046 | 5 | 2.46e-02 | 0 | 0 | 1 | 0 | 0 | 1 |
| 75 | rs187509586 | 1 | 2.54e-02 | 0 | 0 | 0 | 1 | 0 | 1 |
| 41 | rs571956434 | 3 | 2.61e-02 | 1 | 1 | 0 | 0 | 0 | 2 |
| 101 | rs116555012 | 3 | 2.62e-02 | 1 | 0 | 0 | 0 | 0 | 1 |
| 175 | rs74753877 | 8 | 2.70e-02 | 0 | 1 | 0 | 0 | 1 | 2 |

**Table S10:** Selected SNPs, maternal genome.
| Rank | SNP | Chromosome | p-value | Fold |  |  |  |  | Total |
| --- | --- | --- | --- | --- | --- | --- | --- | --- | --- |
|  |  |  |  | 0 | 1 | 2 | 3 | 4 |  |
| 28 | rs7757420 | 6 | 2.79e-02 | 1 | 0 | 1 | 1 | 1 | 4 |
| 216 | rs140613560 | 8 | 2.85e-02 | 1 | 0 | 0 | 0 | 0 | 1 |
| 178 | rs144327778 | 5 | 2.93e-02 | 1 | 0 | 0 | 0 | 0 | 1 |
| 107 | rs117020619 | 16 | 2.94e-02 | 1 | 1 | 0 | 1 | 1 | 4 |
| 33 | rs115905203 | 3 | 3.03e-02 | 0 | 0 | 0 | 1 | 1 | 2 |
| 278 | rs5987938 | 23 | 3.04e-02 | 1 | 0 | 0 | 0 | 0 | 1 |
| 105 | rs191878542 | 6 | 3.48e-02 | 1 | 0 | 0 | 1 | 0 | 2 |
| 9 | rs76677301 | 1 | 3.52e-02 | 0 | 1 | 1 | 1 | 0 | 3 |
| 213 | rs62523909 | 8 | 3.81e-02 | 1 | 0 | 0 | 0 | 0 | 1 |
| 256 | rs528092133 | 16 | 3.86e-02 | 0 | 1 | 0 | 0 | 0 | 1 |
| 257 | rs1076572 | 16 | 3.99e-02 | 0 | 0 | 0 | 1 | 0 | 1 |
| 283 | rs140384340 | 23 | 4.12e-02 | 1 | 0 | 1 | 0 | 1 | 3 |
| 293 | rs139426544 | 23 | 4.25e-02 | 0 | 0 | 1 | 0 | 0 | 1 |
| 208 | rs2495994 | 6 | 4.26e-02 | 0 | 0 | 0 | 0 | 1 | 1 |
| 259 | rs28651817 | 16 | 4.44e-02 | 0 | 0 | 0 | 1 | 0 | 1 |
| 135 | rs73860717 | 3 | 4.48e-02 | 0 | 0 | 0 | 1 | 0 | 1 |

**Table S10:** Selected SNPs, maternal genome.
| Rank | SNP | Chromosome | p-value | Fold |  |  |  |  | Total |
| --- | --- | --- | --- | --- | --- | --- | --- | --- | --- |
|  |  |  |  | 0 | 1 | 2 | 3 | 4 |  |
| 23 | rs13142462 | 4 | 4.55e-02 | 1 | 1 | 0 | 1 | 0 | 3 |
| 212 | rs2495991 | 6 | 4.89e-02 | 0 | 1 | 0 | 0 | 0 | 1 |
| 60 | rs72653572 | 8 | 5.10e-02 | 0 | 1 | 1 | 0 | 1 | 3 |
| 249 | rs77867090 | 9 | 5.12e-02 | 0 | 0 | 1 | 0 | 0 | 1 |
| 250 | rs10120850 | 9 | 5.26e-02 | 0 | 1 | 0 | 0 | 0 | 1 |
| 84 | rs7413155 | 1 | 5.31e-02 | 0 | 0 | 0 | 1 | 0 | 1 |
| 272 | rs10445250 | 17 | 5.34e-02 | 0 | 1 | 0 | 0 | 0 | 1 |
| 36 | rs17276967 | 4 | 5.42e-02 | 1 | 1 | 1 | 0 | 0 | 3 |
| 268 | rs3859250 | 17 | 5.49e-02 | 1 | 0 | 0 | 0 | 0 | 1 |
| 190 | rs12513096 | 4 | 5.65e-02 | 0 | 0 | 0 | 1 | 0 | 1 |
| 171 | rs72716950 | 8 | 5.88e-02 | 1 | 0 | 1 | 0 | 0 | 2 |
| 100 | rs74742427 | 2 | 5.98e-02 | 0 | 0 | 1 | 0 | 0 | 1 |
| 90 | rs77647659 | 8 | 6.21e-02 | 0 | 0 | 1 | 1 | 1 | 3 |
| 70 | rs4655235 | 1 | 6.38e-02 | 0 | 0 | 0 | 0 | 1 | 1 |
| 223 | rs2088663 | 8 | 6.40e-02 | 0 | 0 | 0 | 0 | 1 | 1 |
| 279 | rs138678764 | 17 | 6.92e-02 | 0 | 0 | 0 | 1 | 0 | 1 |

**Table S10:** Selected SNPs, maternal genome.
| Rank | SNP | Chromosome | p-value | Fold |  |  |  |  | Total |
| --- | --- | --- | --- | --- | --- | --- | --- | --- | --- |
|  |  |  |  | 0 | 1 | 2 | 3 | 4 |  |
| 88 | rs13024800 | 2 | 7.46e-02 | 1 | 0 | 0 | 0 | 0 | 1 |
| 49 | rs72806257 | 5 | 7.74e-02 | 0 | 0 | 1 | 1 | 1 | 3 |
| 47 | rs72716936 | 8 | 7.90e-02 | 1 | 1 | 1 | 1 | 1 | 5 |
| 262 | rs117319561 | 16 | 8.64e-02 | 0 | 0 | 1 | 0 | 0 | 1 |
| 2 | rs9854485 | 3 | 8.77e-02 | 1 | 1 | 1 | 1 | 1 | 5 |
| 174 | rs6554337 | 4 | 8.78e-02 | 0 | 0 | 0 | 1 | 0 | 1 |
| 72 | rs35669629 | 5 | 8.94e-02 | 0 | 1 | 0 | 1 | 0 | 2 |
| 134 | rs9882647 | 3 | 9.20e-02 | 0 | 0 | 0 | 1 | 0 | 1 |
| 281 | rs2312056 | 23 | 9.23e-02 | 1 | 0 | 0 | 0 | 0 | 1 |
| 39 | rs114429152 | 3 | 9.26e-02 | 0 | 1 | 0 | 0 | 1 | 2 |
| 189 | rs9501064 | 6 | 9.34e-02 | 1 | 0 | 0 | 0 | 0 | 1 |
| 6 | rs35836191 | 1 | 9.40e-02 | 1 | 0 | 0 | 1 | 1 | 3 |
| 117 | rs188440971 | 3 | 9.54e-02 | 1 | 0 | 0 | 0 | 0 | 1 |
| 182 | rs56229154 | 4 | 9.95e-02 | 0 | 1 | 0 | 0 | 0 | 1 |
| 160 | rs73164735 | 3 | 1.01e-01 | 0 | 0 | 0 | 0 | 1 | 1 |
| 186 | rs11930038 | 4 | 1.02e-01 | 0 | 0 | 0 | 0 | 1 | 1 |

**Table S10:** Selected SNPs, maternal genome.
| Rank | SNP | Chromosome | p-value | Fold |  |  |  |  | Total |
| --- | --- | --- | --- | --- | --- | --- | --- | --- | --- |
|  |  |  |  | 0 | 1 | 2 | 3 | 4 |  |
| 139 | rs2001627 | 3 | 1.05e-01 | 0 | 0 | 0 | 0 | 1 | 1 |
| 14 | rs111541022 | 2 | 1.07e-01 | 0 | 1 | 1 | 0 | 1 | 3 |
| 199 | rs111809046 | 4 | 1.07e-01 | 0 | 1 | 0 | 0 | 0 | 1 |
| 232 | rs79188580 | 8 | 1.09e-01 | 0 | 1 | 0 | 0 | 0 | 1 |
| 137 | rs59303138 | 3 | 1.10e-01 | 0 | 1 | 0 | 0 | 0 | 1 |
| 282 | rs77764993 | 17 | 1.13e-01 | 0 | 0 | 0 | 1 | 0 | 1 |
| 141 | rs528779602 | 3 | 1.16e-01 | 0 | 1 | 0 | 0 | 0 | 1 |
| 127 | rs28654615 | 3 | 1.18e-01 | 1 | 0 | 0 | 0 | 0 | 1 |
| 27 | rs190328567 | 3 | 1.20e-01 | 0 | 1 | 1 | 0 | 1 | 3 |
| 294 | rs75451809 | 20 | 1.29e-01 | 0 | 0 | 0 | 1 | 0 | 1 |
| 104 | rs566709383 | 8 | 1.30e-01 | 0 | 1 | 0 | 0 | 1 | 2 |
| 181 | rs142804514 | 5 | 1.31e-01 | 1 | 0 | 0 | 0 | 0 | 1 |
| 102 | rs62233001 | 3 | 1.33e-01 | 0 | 0 | 0 | 1 | 0 | 1 |
| 290 | rs144545499 | 23 | 1.33e-01 | 1 | 0 | 0 | 0 | 0 | 1 |
| 78 | rs71638874 | 1 | 1.36e-01 | 0 | 0 | 1 | 0 | 0 | 1 |
| 236 | rs112577818 | 17 | 1.36e-01 | 0 | 1 | 1 | 0 | 1 | 3 |

**Table S10:** Selected SNPs, maternal genome.
| Rank | SNP | Chromosome | p-value | Fold |  |  |  |  | Total |
| --- | --- | --- | --- | --- | --- | --- | --- | --- | --- |
|  |  |  |  | 0 | 1 | 2 | 3 | 4 |  |
| 222 | rs116899184 | 8 | 1.40e-01 | 0 | 0 | 1 | 0 | 0 | 1 |
| 246 | rs117311531 | 9 | 1.40e-01 | 0 | 0 | 1 | 0 | 0 | 1 |
| 143 | rs575089268 | 3 | 1.43e-01 | 0 | 1 | 0 | 0 | 0 | 1 |
| 31 | rs17726725 | 2 | 1.45e-01 | 0 | 1 | 0 | 0 | 1 | 2 |
| 241 | rs144243197 | 17 | 1.50e-01 | 1 | 0 | 0 | 0 | 1 | 2 |
| 158 | rs74824193 | 3 | 1.52e-01 | 0 | 1 | 0 | 0 | 0 | 1 |
| 103 | rs1809046 | 3 | 1.54e-01 | 0 | 0 | 1 | 0 | 0 | 1 |
| 19 | rs62273121 | 3 | 1.54e-01 | 1 | 1 | 1 | 0 | 0 | 3 |
| 35 | rs2344719 | 3 | 1.56e-01 | 1 | 1 | 0 | 0 | 0 | 2 |
| 234 | rs186486331 | 8 | 1.58e-01 | 0 | 0 | 0 | 1 | 0 | 1 |
| 93 | rs145412470 | 3 | 1.59e-01 | 1 | 0 | 0 | 0 | 0 | 1 |
| 80 | rs115120397 | 1 | 1.60e-01 | 0 | 1 | 0 | 0 | 0 | 1 |
| 53 | rs72718778 | 8 | 1.60e-01 | 1 | 1 | 1 | 1 | 1 | 5 |
| 274 | rs2024082 | 17 | 1.61e-01 | 0 | 0 | 1 | 0 | 0 | 1 |
| 82 | rs12041540 | 1 | 1.66e-01 | 0 | 0 | 0 | 0 | 1 | 1 |
| 267 | rs117565066 | 16 | 1.72e-01 | 0 | 0 | 0 | 1 | 0 | 1 |

**Table S10:** Selected SNPs, maternal genome.
| Rank | SNP | Chromosome | p-value | Fold |  |  |  |  | Total |
| --- | --- | --- | --- | --- | --- | --- | --- | --- | --- |
|  |  |  |  | 0 | 1 | 2 | 3 | 4 |  |
| 131 | rs138513400 | 3 | 1.77e-01 | 0 | 0 | 0 | 0 | 1 | 1 |
| 98 | rs188291622 | 2 | 1.88e-01 | 0 | 0 | 0 | 0 | 1 | 1 |
| 210 | rs9313828 | 5 | 1.91e-01 | 0 | 0 | 0 | 1 | 0 | 1 |
| 240 | rs67259800 | 8 | 1.92e-01 | 0 | 0 | 1 | 0 | 0 | 1 |
| 254 | rs182204471 | 16 | 1.93e-01 | 0 | 0 | 0 | 0 | 1 | 1 |
| 264 | rs75379427 | 20 | 2.00e-01 | 0 | 1 | 0 | 1 | 1 | 3 |
| 54 | rs34744948 | 6 | 2.01e-01 | 1 | 0 | 0 | 1 | 1 | 3 |
| 221 | rs56942423 | 17 | 2.04e-01 | 1 | 1 | 1 | 1 | 1 | 5 |
| 30 | rs115628998 | 4 | 2.06e-01 | 1 | 1 | 1 | 0 | 0 | 3 |
| 155 | rs57771301 | 3 | 2.08e-01 | 0 | 0 | 0 | 1 | 0 | 1 |
| 109 | rs76684625 | 3 | 2.08e-01 | 0 | 0 | 0 | 1 | 0 | 1 |
| 17 | rs78120455 | 3 | 2.13e-01 | 0 | 1 | 0 | 1 | 1 | 3 |
| 195 | rs149713782 | 9 | 2.14e-01 | 1 | 0 | 0 | 1 | 0 | 2 |
| 118 | rs146272610 | 3 | 2.16e-01 | 0 | 0 | 0 | 1 | 0 | 1 |
| 87 | rs80212941 | 1 | 2.19e-01 | 0 | 0 | 0 | 1 | 0 | 1 |
| 7 | rs149527606 | 4 | 2.24e-01 | 1 | 1 | 1 | 1 | 1 | 5 |

**Table S10:** Selected SNPs, maternal genome.
| Rank | SNP | Chromosome | p-value | Fold |  |  |  |  | Total |
| --- | --- | --- | --- | --- | --- | --- | --- | --- | --- |
|  |  |  |  | 0 | 1 | 2 | 3 | 4 |  |
| 113 | rs184519786 | 3 | 2.24e-01 | 0 | 0 | 0 | 1 | 0 | 1 |
| 110 | rs2332238 | 3 | 2.26e-01 | 1 | 0 | 0 | 0 | 0 | 1 |
| 46 | rs76069554 | 5 | 2.28e-01 | 1 | 0 | 0 | 1 | 1 | 3 |
| 11 | rs140224603 | 1 | 2.34e-01 | 1 | 1 | 0 | 0 | 1 | 3 |
| 179 | rs1351212 | 4 | 2.35e-01 | 0 | 1 | 0 | 0 | 0 | 1 |
| 52 | rs34841490 | 3 | 2.36e-01 | 0 | 0 | 1 | 1 | 0 | 2 |
| 154 | rs139090679 | 16 | 2.42e-01 | 1 | 1 | 1 | 0 | 1 | 4 |
| 51 | rs73203403 | 3 | 2.43e-01 | 0 | 0 | 1 | 0 | 1 | 2 |
| 265 | rs59451439 | 17 | 2.47e-01 | 0 | 0 | 0 | 0 | 1 | 1 |
| 202 | rs34658030 | 5 | 2.49e-01 | 0 | 1 | 0 | 0 | 0 | 1 |
| 215 | rs9267484 | 6 | 2.57e-01 | 0 | 0 | 1 | 0 | 0 | 1 |
| 76 | rs139831675 | 1 | 2.58e-01 | 0 | 0 | 0 | 0 | 1 | 1 |
| 43 | rs1965290 | 3 | 2.68e-01 | 1 | 0 | 0 | 0 | 1 | 2 |
| 273 | rs117941433 | 17 | 2.70e-01 | 0 | 0 | 0 | 0 | 1 | 1 |
| 280 | rs146923460 | 17 | 2.71e-01 | 0 | 0 | 0 | 1 | 0 | 1 |
| 115 | rs75779179 | 3 | 2.72e-01 | 0 | 0 | 1 | 0 | 0 | 1 |

**Table S10:** Selected SNPs, maternal genome.
| Rank | SNP | Chromosome | p-value | Fold |  |  |  |  | Total |
| --- | --- | --- | --- | --- | --- | --- | --- | --- | --- |
|  |  |  |  | 0 | 1 | 2 | 3 | 4 |  |
| 198 | rs138144869 | 16 | 2.76e-01 | 0 | 0 | 1 | 1 | 1 | 3 |
| 289 | rs80019664 | 20 | 2.84e-01 | 0 | 0 | 0 | 0 | 1 | 1 |
| 144 | rs9872573 | 3 | 2.89e-01 | 0 | 0 | 0 | 0 | 1 | 1 |
| 81 | rs112548458 | 1 | 2.92e-01 | 1 | 0 | 0 | 0 | 0 | 1 |
| 85 | rs55921738 | 1 | 2.95e-01 | 0 | 0 | 1 | 0 | 0 | 1 |
| 287 | rs117114983 | 20 | 2.96e-01 | 0 | 0 | 1 | 0 | 0 | 1 |
| 245 | rs584190 | 9 | 3.02e-01 | 0 | 1 | 0 | 0 | 0 | 1 |
| 196 | rs147977333 | 4 | 3.06e-01 | 0 | 0 | 0 | 0 | 1 | 1 |
| 214 | rs111500568 | 16 | 3.42e-01 | 1 | 1 | 0 | 1 | 0 | 3 |
| 225 | rs78372378 | 6 | 3.43e-01 | 0 | 0 | 0 | 1 | 0 | 1 |
| 57 | rs62574455 | 9 | 3.46e-01 | 1 | 1 | 1 | 1 | 1 | 5 |
| 65 | rs7654438 | 4 | 3.46e-01 | 1 | 0 | 1 | 0 | 0 | 2 |
| 296 | rs5930616 | 23 | 3.51e-01 | 1 | 1 | 0 | 0 | 0 | 2 |
| 136 | rs78416779 | 8 | 3.54e-01 | 0 | 1 | 1 | 0 | 0 | 2 |
| 67 | rs115439992 | 5 | 3.54e-01 | 1 | 0 | 1 | 0 | 0 | 2 |
| 261 | rs129986 | 16 | 3.55e-01 | 0 | 0 | 0 | 1 | 0 | 1 |

**Table S10:** Selected SNPs, maternal genome.
| Rank | SNP | Chromosome | p-value | Fold |  |  |  |  | Total |
| --- | --- | --- | --- | --- | --- | --- | --- | --- | --- |
|  |  |  |  | 0 | 1 | 2 | 3 | 4 |  |
| 277 | rs74656135 | 17 | 3.57e-01 | 0 | 0 | 0 | 0 | 1 | 1 |
| 44 | rs138176253 | 3 | 3.58e-01 | 1 | 0 | 1 | 0 | 0 | 2 |
| 288 | rs149928287 | 23 | 3.62e-01 | 1 | 0 | 0 | 0 | 0 | 1 |
| 142 | rs139321243 | 3 | 3.62e-01 | 0 | 0 | 1 | 0 | 0 | 1 |
| 167 | rs147255257 | 16 | 3.62e-01 | 1 | 1 | 1 | 1 | 0 | 4 |
| 286 | rs76604150 | 20 | 3.62e-01 | 0 | 0 | 0 | 0 | 1 | 1 |
| 8 | rs150794748 | 1 | 3.62e-01 | 1 | 1 | 0 | 1 | 0 | 3 |
| 284 | rs2001124 | 23 | 3.66e-01 | 1 | 0 | 0 | 0 | 0 | 1 |
| 270 | rs71364197 | 17 | 3.66e-01 | 0 | 1 | 0 | 0 | 0 | 1 |
| 211 | rs129994 | 16 | 3.67e-01 | 0 | 0 | 1 | 1 | 1 | 3 |
| 235 | rs141062087 | 8 | 3.68e-01 | 0 | 0 | 0 | 0 | 1 | 1 |
| 218 | rs438475 | 6 | 3.69e-01 | 0 | 0 | 0 | 1 | 0 | 1 |
| 229 | rs2259214 | 16 | 3.81e-01 | 0 | 0 | 1 | 0 | 1 | 2 |
| 128 | rs145683289 | 8 | 3.83e-01 | 1 | 1 | 0 | 0 | 0 | 2 |
| 122 | rs13066825 | 3 | 3.87e-01 | 0 | 0 | 0 | 0 | 1 | 1 |
| 184 | rs4615153 | 4 | 3.87e-01 | 0 | 0 | 0 | 1 | 0 | 1 |

**Table S10:** Selected SNPs, maternal genome.
| Rank | SNP | Chromosome | p-value | Fold |  |  |  |  | Total |
| --- | --- | --- | --- | --- | --- | --- | --- | --- | --- |
|  |  |  |  | 0 | 1 | 2 | 3 | 4 |  |
| 253 | rs79191736 | 16 | 4.04e-01 | 1 | 0 | 0 | 0 | 0 | 1 |
| 169 | rs7459568 | 8 | 4.13e-01 | 0 | 1 | 1 | 0 | 0 | 2 |
| 153 | rs73240547 | 4 | 4.22e-01 | 1 | 0 | 0 | 0 | 0 | 1 |
| 248 | rs77822078 | 17 | 4.23e-01 | 1 | 1 | 0 | 0 | 0 | 2 |
| 176 | rs56383248 | 4 | 4.26e-01 | 0 | 0 | 0 | 0 | 1 | 1 |
| 255 | rs113879805 | 16 | 4.32e-01 | 1 | 0 | 0 | 0 | 0 | 1 |
| 266 | rs139669705 | 17 | 4.33e-01 | 0 | 1 | 1 | 0 | 0 | 2 |
| 15 | rs7636691 | 3 | 4.34e-01 | 0 | 1 | 0 | 1 | 1 | 3 |
| 263 | rs74252988 | 17 | 4.40e-01 | 0 | 1 | 0 | 0 | 0 | 1 |
| 161 | rs146754796 | 16 | 4.40e-01 | 1 | 1 | 1 | 1 | 1 | 5 |
| 5 | rs140529843 | 1 | 4.43e-01 | 1 | 1 | 1 | 0 | 1 | 4 |
| 170 | rs112586832 | 4 | 4.45e-01 | 0 | 0 | 0 | 1 | 0 | 1 |
| 185 | rs8177804 | 9 | 4.50e-01 | 1 | 0 | 0 | 1 | 0 | 2 |
| 276 | rs142167053 | 20 | 4.53e-01 | 1 | 0 | 0 | 0 | 0 | 1 |
| 230 | rs16897941 | 8 | 4.53e-01 | 0 | 0 | 1 | 0 | 0 | 1 |
| 209 | rs2451375 | 6 | 4.71e-01 | 1 | 0 | 0 | 0 | 0 | 1 |

**Table S10:** Selected SNPs, maternal genome.
| Rank | SNP | Chromosome | p-value | Fold |  |  |  |  | Total |
| --- | --- | --- | --- | --- | --- | --- | --- | --- | --- |
|  |  |  |  | 0 | 1 | 2 | 3 | 4 |  |
| 197 | rs189769609 | 4 | 4.73e-01 | 0 | 0 | 0 | 1 | 0 | 1 |
| 123 | rs58563555 | 3 | 4.75e-01 | 1 | 0 | 0 | 0 | 0 | 1 |
| 91 | rs189442736 | 2 | 4.76e-01 | 0 | 0 | 1 | 0 | 0 | 1 |
| 242 | rs2153838 | 9 | 4.76e-01 | 0 | 1 | 0 | 0 | 0 | 1 |
| 116 | rs12490230 | 3 | 4.82e-01 | 0 | 0 | 1 | 0 | 0 | 1 |
| 163 | rs78012168 | 3 | 4.86e-01 | 0 | 0 | 0 | 0 | 1 | 1 |
| 159 | rs74770666 | 3 | 4.86e-01 | 0 | 0 | 1 | 0 | 0 | 1 |
| 74 | rs61769198 | 1 | 4.89e-01 | 0 | 0 | 0 | 0 | 1 | 1 |
| 55 | rs7643005 | 3 | 5.01e-01 | 0 | 0 | 1 | 1 | 0 | 2 |
| 99 | rs2080229 | 2 | 5.01e-01 | 0 | 1 | 0 | 0 | 0 | 1 |
| 285 | rs138458916 | 20 | 5.02e-01 | 0 | 1 | 0 | 0 | 0 | 1 |
| 239 | rs150392264 | 17 | 5.10e-01 | 1 | 1 | 0 | 0 | 0 | 2 |
| 4 | rs67874885 | 1 | 5.11e-01 | 1 | 1 | 1 | 1 | 0 | 4 |
| 231 | rs72766568 | 16 | 5.25e-01 | 0 | 1 | 0 | 0 | 1 | 2 |
| 77 | rs2075995 | 1 | 5.26e-01 | 1 | 0 | 0 | 0 | 0 | 1 |
| 83 | rs169695 | 1 | 5.31e-01 | 0 | 0 | 1 | 0 | 0 | 1 |

**Table S10:** Selected SNPs, maternal genome.
| Rank | SNP | Chromosome | p-value | Fold |  |  |  |  | Total |
| --- | --- | --- | --- | --- | --- | --- | --- | --- | --- |
|  |  |  |  | 0 | 1 | 2 | 3 | 4 |  |
| 106 | rs12469594 | 2 | 5.35e-01 | 0 | 0 | 0 | 0 | 1 | 1 |
| 38 | rs77169019 | 6 | 5.36e-01 | 0 | 1 | 1 | 1 | 1 | 4 |
| 97 | rs6723410 | 2 | 5.39e-01 | 0 | 1 | 0 | 0 | 0 | 1 |
| 119 | rs73337567 | 8 | 5.58e-01 | 0 | 1 | 0 | 0 | 1 | 2 |
| 206 | rs67101146 | 5 | 5.62e-01 | 0 | 0 | 0 | 1 | 0 | 1 |
| 24 | rs76035212 | 1 | 5.79e-01 | 0 | 0 | 1 | 0 | 1 | 2 |
| 162 | rs28489443 | 4 | 5.79e-01 | 0 | 0 | 0 | 1 | 0 | 1 |
| 126 | rs147137688 | 8 | 5.81e-01 | 0 | 1 | 0 | 0 | 1 | 2 |
| 173 | rs143940877 | 4 | 5.86e-01 | 0 | 1 | 0 | 0 | 0 | 1 |
| 89 | rs75434492 | 1 | 5.86e-01 | 0 | 0 | 0 | 0 | 1 | 1 |
| 219 | rs78535051 | 8 | 5.87e-01 | 0 | 0 | 0 | 0 | 1 | 1 |
| 164 | rs2581628 | 3 | 5.89e-01 | 0 | 0 | 1 | 0 | 0 | 1 |
| 86 | rs80089098 | 1 | 5.89e-01 | 0 | 0 | 0 | 0 | 1 | 1 |
| 69 | rs7538997 | 1 | 5.97e-01 | 0 | 0 | 1 | 0 | 0 | 1 |
| 22 | rs72659201 | 1 | 6.13e-01 | 0 | 1 | 0 | 1 | 0 | 2 |
| 192 | rs184291275 | 4 | 6.15e-01 | 0 | 0 | 0 | 0 | 1 | 1 |

**Table S10:** Selected SNPs, maternal genome.
| Rank | SNP | Chromosome | p-value | Fold |  |  |  |  | Total |
| --- | --- | --- | --- | --- | --- | --- | --- | --- | --- |
|  |  |  |  | 0 | 1 | 2 | 3 | 4 |  |
| 217 | rs117000870 | 8 | 6.20e-01 | 1 | 0 | 0 | 0 | 0 | 1 |
| 260 | rs148078592 | 17 | 6.26e-01 | 1 | 0 | 0 | 0 | 0 | 1 |
| 226 | rs186536363 | 8 | 6.43e-01 | 0 | 0 | 1 | 0 | 0 | 1 |
| 48 | rs28537590 | 8 | 6.64e-01 | 1 | 1 | 1 | 0 | 1 | 4 |
| 238 | rs72720099 | 8 | 6.65e-01 | 0 | 0 | 0 | 1 | 0 | 1 |
| 50 | rs78216422 | 3 | 6.66e-01 | 1 | 0 | 0 | 0 | 1 | 2 |
| 292 | rs186125715 | 23 | 6.68e-01 | 0 | 1 | 0 | 0 | 0 | 1 |
| 40 | rs79726092 | 3 | 6.68e-01 | 0 | 0 | 1 | 0 | 1 | 2 |
| 233 | rs1905045 | 8 | 6.72e-01 | 0 | 0 | 0 | 1 | 0 | 1 |
| 108 | rs116415411 | 3 | 6.73e-01 | 0 | 0 | 1 | 0 | 0 | 1 |
| 130 | rs116182279 | 3 | 6.90e-01 | 0 | 1 | 0 | 0 | 0 | 1 |
| 111 | rs829185 | 3 | 7.02e-01 | 0 | 0 | 0 | 1 | 0 | 1 |
| 224 | rs117622394 | 8 | 7.06e-01 | 1 | 0 | 0 | 0 | 0 | 1 |
| 32 | rs13084063 | 3 | 7.15e-01 | 1 | 1 | 0 | 0 | 0 | 2 |
| 132 | rs1028259 | 8 | 7.23e-01 | 0 | 1 | 0 | 0 | 1 | 2 |
| 168 | rs116152825 | 4 | 7.58e-01 | 1 | 0 | 0 | 0 | 0 | 1 |

**Table S10:** Selected SNPs, maternal genome.
| Rank | SNP | Chromosome | p-value | Fold |  |  |  |  | Total |
| --- | --- | --- | --- | --- | --- | --- | --- | --- | --- |
|  |  |  |  | 0 | 1 | 2 | 3 | 4 |  |
| 269 | rs73510369 | 16 | 7.63e-01 | 0 | 0 | 0 | 1 | 0 | 1 |
| 275 | rs758434 | 17 | 7.72e-01 | 0 | 0 | 0 | 1 | 0 | 1 |
| 220 | rs72709825 | 8 | 7.93e-01 | 1 | 0 | 0 | 0 | 0 | 1 |
| 26 | rs17083076 | 4 | 7.99e-01 | 1 | 0 | 1 | 0 | 1 | 3 |
| 148 | rs79835427 | 3 | 8.16e-01 | 0 | 1 | 0 | 0 | 0 | 1 |
| 271 | rs116491757 | 20 | 8.38e-01 | 1 | 0 | 0 | 0 | 0 | 1 |
| 68 | rs76266717 | 1 | 8.50e-01 | 0 | 0 | 0 | 1 | 0 | 1 |
| 243 | rs116864430 | 9 | 8.63e-01 | 0 | 0 | 1 | 0 | 0 | 1 |
| 66 | rs30142 | 5 | 8.63e-01 | 1 | 0 | 1 | 0 | 0 | 2 |
| 227 | rs2495980 | 6 | 8.64e-01 | 0 | 0 | 0 | 1 | 0 | 1 |
| 120 | rs34407651 | 3 | 8.81e-01 | 0 | 1 | 0 | 0 | 0 | 1 |
| 145 | rs34566348 | 3 | 8.91e-01 | 0 | 1 | 0 | 0 | 0 | 1 |
| 0 | rs41302778 | 1 | 8.94e-01 | 1 | 1 | 1 | 1 | 1 | 5 |
| 138 | rs74669781 | 3 | 9.06e-01 | 0 | 1 | 0 | 0 | 0 | 1 |
| 237 | rs4870922 | 8 | 9.08e-01 | 0 | 0 | 1 | 0 | 0 | 1 |
| 112 | rs75037852 | 3 | 9.11e-01 | 0 | 0 | 1 | 0 | 0 | 1 |

**Table S10:** Selected SNPs, maternal genome.
| Rank | SNP | Chromosome | p-value | Fold |  |  |  |  | Total |
| --- | --- | --- | --- | --- | --- | --- | --- | --- | --- |
|  |  |  |  | 0 | 1 | 2 | 3 | 4 |  |
| 151 | rs55854104 | 3 | 9.12e-01 | 0 | 0 | 1 | 0 | 0 | 1 |
| 157 | rs138490397 | 8 | 9.37e-01 | 0 | 1 | 1 | 0 | 0 | 2 |
| 12 | rs74919188 | 2 | 9.56e-01 | 1 | 1 | 0 | 1 | 0 | 3 |
| 252 | rs146180973 | 9 | 9.65e-01 | 0 | 0 | 1 | 0 | 0 | 1 |
| 94 | rs116265088 | 2 | 9.73e-01 | 0 | 0 | 1 | 0 | 0 | 1 |
| 165 | rs1531290 | 4 | 9.79e-01 | 0 | 0 | 0 | 1 | 0 | 1 |
| 247 | rs13287637 | 9 | 9.81e-01 | 0 | 0 | 0 | 1 | 0 | 1 |
| 258 | rs146916265 | 9 | 9.89e-01 | 0 | 0 | 1 | 0 | 0 | 1 |

**Table S11:** Selected SNPs, fetal genome. List of the SNPs in the Selected subset for the fetal genome, sorted by p-value. Rank is calculated over all folds and the total number of folds where the SNP is included is listed under Total.

| Rank | SNP | Chromosome | p-value | Fold |  |  |  |  | Total |
| --- | --- | --- | --- | --- | --- | --- | --- | --- | --- |
|  |  |  |  | 0 | 1 | 2 | 3 | 4 |  |
| 167 | rs113828443 | 4 | 1.72e-09 | 1 | 0 | 0 | 0 | 0 | 1 |
| 59 | rs116449362 | 4 | 5.46e-07 | 1 | 1 | 0 | 0 | 0 | 2 |
| 178 | rs2269712 | 6 | 8.06e-07 | 0 | 0 | 0 | 0 | 1 | 1 |

**Table S11:** Selected SNPs, fetal genome.
| Rank | SNP | Chromosome | p-value | Fold |  |  |  |  | Total |
| --- | --- | --- | --- | --- | --- | --- | --- | --- | --- |
|  |  |  |  | 0 | 1 | 2 | 3 | 4 |  |
| 57 | rs58218495 | 3 | 1.09e-06 | 1 | 0 | 1 | 0 | 0 | 2 |
| 84 | rs150703020 | 1 | 6.03e-06 | 1 | 0 | 0 | 0 | 0 | 1 |
| 88 | rs115272033 | 6 | 1.04e-05 | 1 | 0 | 0 | 0 | 1 | 2 |
| 176 | rs17634544 | 5 | 1.20e-04 | 0 | 0 | 0 | 0 | 1 | 1 |
| 81 | rs184415473 | 1 | 1.33e-04 | 0 | 0 | 1 | 0 | 0 | 1 |
| 134 | rs7626926 | 3 | 1.34e-04 | 0 | 1 | 0 | 0 | 0 | 1 |
| 10 | rs13061037 | 3 | 4.47e-04 | 1 | 1 | 0 | 1 | 1 | 4 |
| 45 | rs206018 | 6 | 5.30e-04 | 0 | 1 | 0 | 1 | 1 | 3 |
| 76 | rs10917042 | 1 | 6.04e-04 | 0 | 0 | 0 | 0 | 1 | 1 |
| 41 | rs2844543 | 6 | 6.77e-04 | 0 | 1 | 1 | 0 | 1 | 3 |
| 40 | rs138906059 | 8 | 6.84e-04 | 1 | 1 | 0 | 1 | 1 | 4 |
| 182 | rs2853915 | 6 | 9.13e-04 | 0 | 0 | 1 | 0 | 0 | 1 |
| 107 | rs11707570 | 3 | 1.08e-03 | 0 | 0 | 0 | 1 | 0 | 1 |
| 38 | rs12681283 | 8 | 1.12e-03 | 0 | 1 | 1 | 1 | 1 | 4 |
| 258 | rs2427493 | 20 | 1.82e-03 | 0 | 1 | 0 | 0 | 0 | 1 |
| 154 | rs2332796 | 4 | 2.01e-03 | 0 | 1 | 0 | 0 | 0 | 1 |

**Table S11:** Selected SNPs, fetal genome.
| Rank | SNP | Chromosome | p-value | Fold |  |  |  |  | Total |
| --- | --- | --- | --- | --- | --- | --- | --- | --- | --- |
|  |  |  |  | 0 | 1 | 2 | 3 | 4 |  |
| 143 | rs11133345 | 4 | 2.27e-03 | 0 | 1 | 0 | 0 | 0 | 1 |
| 242 | rs117169569 | 17 | 2.62e-03 | 0 | 1 | 0 | 0 | 0 | 1 |
| 93 | rs2857702 | 6 | 2.92e-03 | 0 | 0 | 1 | 0 | 1 | 2 |
| 157 | rs190596665 | 5 | 5.25e-03 | 0 | 0 | 0 | 1 | 0 | 1 |
| 117 | rs6803805 | 3 | 6.60e-03 | 0 | 1 | 0 | 0 | 0 | 1 |
| 77 | rs11799887 | 1 | 6.77e-03 | 0 | 0 | 1 | 0 | 0 | 1 |
| 173 | rs35483114 | 5 | 6.93e-03 | 0 | 0 | 0 | 0 | 1 | 1 |
| 196 | rs7821472 | 8 | 7.21e-03 | 0 | 0 | 0 | 1 | 0 | 1 |
| 201 | rs210210 | 6 | 7.28e-03 | 0 | 0 | 1 | 0 | 0 | 1 |
| 64 | rs149863069 | 9 | 7.52e-03 | 1 | 1 | 1 | 0 | 1 | 4 |
| 61 | rs4547776 | 4 | 8.44e-03 | 0 | 1 | 1 | 0 | 0 | 2 |
| 136 | rs12496940 | 3 | 8.68e-03 | 1 | 0 | 0 | 0 | 0 | 1 |
| 55 | rs1978600 | 3 | 1.11e-02 | 1 | 0 | 0 | 0 | 1 | 2 |
| 166 | rs76334428 | 4 | 1.16e-02 | 0 | 0 | 1 | 0 | 0 | 1 |
| 198 | rs184003 | 6 | 1.29e-02 | 0 | 0 | 0 | 0 | 1 | 1 |
| 9 | rs9854729 | 3 | 1.29e-02 | 1 | 1 | 1 | 0 | 1 | 4 |

**Table S11:** Selected SNPs, fetal genome.
| Rank | SNP | Chromosome | p-value | Fold |  |  |  |  | Total |
| --- | --- | --- | --- | --- | --- | --- | --- | --- | --- |
|  |  |  |  | 0 | 1 | 2 | 3 | 4 |  |
| 7 | rs9917792 | 3 | 1.34e-02 | 1 | 1 | 1 | 1 | 0 | 4 |
| 220 | rs75217590 | 8 | 1.37e-02 | 0 | 0 | 1 | 0 | 0 | 1 |
| 267 | rs188603317 | 23 | 1.37e-02 | 0 | 1 | 0 | 0 | 0 | 1 |
| 49 | rs2844463 | 6 | 1.37e-02 | 0 | 0 | 1 | 1 | 1 | 3 |
| 231 | rs9302815 | 16 | 1.51e-02 | 0 | 0 | 0 | 1 | 0 | 1 |
| 72 | rs2844580 | 6 | 1.61e-02 | 0 | 1 | 0 | 0 | 1 | 2 |
| 44 | rs56834107 | 3 | 1.71e-02 | 0 | 1 | 1 | 0 | 0 | 2 |
| 226 | rs78336371 | 17 | 1.74e-02 | 1 | 1 | 1 | 0 | 0 | 3 |
| 24 | rs118027082 | 8 | 1.81e-02 | 1 | 1 | 1 | 1 | 1 | 5 |
| 114 | rs33914662 | 3 | 1.95e-02 | 0 | 0 | 0 | 1 | 0 | 1 |
| 113 | rs9825582 | 3 | 2.01e-02 | 0 | 0 | 0 | 0 | 1 | 1 |
| 282 | rs2119661 | 23 | 2.01e-02 | 1 | 0 | 0 | 0 | 1 | 2 |
| 214 | rs143620094 | 8 | 2.06e-02 | 0 | 0 | 1 | 0 | 0 | 1 |
| 62 | rs74746413 | 5 | 2.06e-02 | 0 | 1 | 0 | 1 | 0 | 2 |
| 152 | rs112199365 | 4 | 2.14e-02 | 0 | 0 | 0 | 1 | 0 | 1 |
| 23 | rs78793891 | 3 | 2.17e-02 | 1 | 0 | 1 | 0 | 1 | 3 |

**Table S11:** Selected SNPs, fetal genome.
| Rank | SNP | Chromosome | p-value | Fold |  |  |  |  | Total |
| --- | --- | --- | --- | --- | --- | --- | --- | --- | --- |
|  |  |  |  | 0 | 1 | 2 | 3 | 4 |  |
| 179 | rs2269476 | 6 | 2.21e-02 | 1 | 0 | 0 | 0 | 0 | 1 |
| 99 | rs150002738 | 2 | 2.40e-02 | 0 | 0 | 0 | 1 | 0 | 1 |
| 130 | rs112545155 | 8 | 2.47e-02 | 1 | 1 | 0 | 0 | 0 | 2 |
| 145 | rs143166647 | 9 | 2.48e-02 | 1 | 0 | 0 | 1 | 0 | 2 |
| 89 | rs111905599 | 3 | 2.53e-02 | 0 | 0 | 1 | 0 | 0 | 1 |
| 244 | rs3915568 | 20 | 2.54e-02 | 0 | 1 | 1 | 0 | 0 | 2 |
| 18 | rs9264931 | 6 | 2.55e-02 | 1 | 0 | 1 | 1 | 1 | 4 |
| 110 | rs6764544 | 3 | 2.60e-02 | 0 | 0 | 0 | 1 | 0 | 1 |
| 191 | rs2981106 | 8 | 2.64e-02 | 1 | 0 | 0 | 0 | 0 | 1 |
| 163 | rs150479458 | 3 | 2.73e-02 | 0 | 0 | 0 | 0 | 1 | 1 |
| 21 | rs7757420 | 6 | 2.79e-02 | 1 | 0 | 1 | 1 | 1 | 4 |
| 144 | rs75742486 | 3 | 2.92e-02 | 0 | 0 | 1 | 0 | 0 | 1 |
| 35 | rs114687191 | 3 | 2.98e-02 | 0 | 1 | 1 | 0 | 0 | 2 |
| 254 | rs149177736 | 20 | 3.00e-02 | 0 | 0 | 0 | 1 | 1 | 2 |
| 271 | rs5987938 | 23 | 3.04e-02 | 1 | 0 | 0 | 0 | 0 | 1 |
| 227 | rs6981534 | 8 | 3.08e-02 | 0 | 0 | 1 | 0 | 0 | 1 |

**Table S11:** Selected SNPs, fetal genome.
| Rank | SNP | Chromosome | p-value | Fold |  |  |  |  | Total |
| --- | --- | --- | --- | --- | --- | --- | --- | --- | --- |
|  |  |  |  | 0 | 1 | 2 | 3 | 4 |  |
| 80 | rs145607311 | 1 | 3.16e-02 | 0 | 0 | 0 | 0 | 1 | 1 |
| 235 | rs41278663 | 9 | 3.23e-02 | 0 | 0 | 1 | 0 | 0 | 1 |
| 149 | rs7184446 | 16 | 3.54e-02 | 1 | 1 | 1 | 0 | 0 | 3 |
| 229 | rs72714706 | 8 | 3.63e-02 | 0 | 0 | 1 | 0 | 0 | 1 |
| 232 | rs528092133 | 16 | 3.86e-02 | 0 | 0 | 0 | 1 | 0 | 1 |
| 3 | rs62233032 | 3 | 4.11e-02 | 1 | 1 | 1 | 1 | 1 | 5 |
| 224 | rs9926873 | 16 | 4.16e-02 | 0 | 1 | 0 | 0 | 0 | 1 |
| 261 | rs2281566 | 20 | 4.21e-02 | 0 | 1 | 0 | 0 | 0 | 1 |
| 36 | rs13001436 | 2 | 4.27e-02 | 1 | 0 | 0 | 1 | 0 | 2 |
| 17 | rs2495991 | 6 | 4.89e-02 | 1 | 1 | 1 | 1 | 1 | 5 |
| 168 | rs1991493 | 5 | 5.28e-02 | 0 | 1 | 0 | 0 | 0 | 1 |
| 248 | rs3859250 | 17 | 5.49e-02 | 1 | 0 | 0 | 0 | 0 | 1 |
| 102 | rs3773252 | 3 | 5.82e-02 | 1 | 0 | 0 | 0 | 0 | 1 |
| 65 | rs192738111 | 5 | 5.98e-02 | 0 | 1 | 0 | 1 | 0 | 2 |
| 155 | rs1551066 | 3 | 6.11e-02 | 0 | 0 | 0 | 0 | 1 | 1 |
| 204 | rs17138385 | 16 | 6.25e-02 | 1 | 0 | 0 | 0 | 1 | 2 |

**Table S11:** Selected SNPs, fetal genome.
| Rank | SNP | Chromosome | p-value | Fold |  |  |  |  | Total |
| --- | --- | --- | --- | --- | --- | --- | --- | --- | --- |
|  |  |  |  | 0 | 1 | 2 | 3 | 4 |  |
| 194 | rs12544430 | 8 | 6.28e-02 | 1 | 0 | 0 | 0 | 0 | 1 |
| 33 | rs76295843 | 2 | 6.57e-02 | 1 | 0 | 0 | 1 | 0 | 2 |
| 133 | rs79178498 | 16 | 6.62e-02 | 1 | 0 | 1 | 0 | 1 | 3 |
| 262 | rs187800701 | 20 | 6.72e-02 | 0 | 1 | 0 | 0 | 0 | 1 |
| 126 | rs13098447 | 3 | 6.83e-02 | 0 | 1 | 0 | 0 | 0 | 1 |
| 251 | rs8076936 | 17 | 6.85e-02 | 0 | 0 | 0 | 1 | 0 | 1 |
| 79 | rs71648543 | 1 | 7.21e-02 | 0 | 0 | 1 | 0 | 0 | 1 |
| 27 | rs4660489 | 1 | 7.26e-02 | 0 | 0 | 0 | 1 | 1 | 2 |
| 278 | rs62219717 | 20 | 7.48e-02 | 0 | 0 | 1 | 0 | 0 | 1 |
| 5 | rs145590853 | 3 | 7.76e-02 | 1 | 1 | 1 | 1 | 1 | 5 |
| 96 | rs13430504 | 2 | 8.57e-02 | 0 | 0 | 0 | 0 | 1 | 1 |
| 112 | rs138048566 | 3 | 8.65e-02 | 0 | 0 | 1 | 0 | 0 | 1 |
| 192 | rs115277907 | 6 | 8.69e-02 | 0 | 0 | 1 | 0 | 0 | 1 |
| 127 | rs4026934 | 3 | 8.74e-02 | 0 | 0 | 0 | 1 | 0 | 1 |
| 125 | rs10956169 | 8 | 8.75e-02 | 0 | 1 | 0 | 1 | 0 | 2 |
| 280 | rs138995825 | 23 | 8.79e-02 | 1 | 0 | 0 | 1 | 0 | 2 |

**Table S11:** Selected SNPs, fetal genome.
| Rank | SNP | Chromosome | p-value | Fold |  |  |  |  | Total |
| --- | --- | --- | --- | --- | --- | --- | --- | --- | --- |
|  |  |  |  | 0 | 1 | 2 | 3 | 4 |  |
| 175 | rs145219649 | 5 | 9.07e-02 | 0 | 0 | 1 | 0 | 0 | 1 |
| 43 | rs9882647 | 3 | 9.20e-02 | 0 | 0 | 0 | 1 | 1 | 2 |
| 247 | rs2312056 | 23 | 9.23e-02 | 1 | 0 | 1 | 1 | 1 | 4 |
| 252 | rs140986549 | 17 | 9.34e-02 | 1 | 0 | 0 | 0 | 0 | 1 |
| 170 | rs72694227 | 4 | 9.39e-02 | 0 | 0 | 0 | 0 | 1 | 1 |
| 13 | rs188440971 | 3 | 9.54e-02 | 1 | 1 | 1 | 0 | 0 | 3 |
| 140 | rs9825227 | 3 | 9.61e-02 | 0 | 0 | 0 | 0 | 1 | 1 |
| 237 | rs12448193 | 16 | 9.87e-02 | 0 | 1 | 0 | 0 | 0 | 1 |
| 71 | rs72713213 | 8 | 1.00e-01 | 1 | 1 | 0 | 1 | 0 | 3 |
| 39 | rs117868519 | 8 | 1.01e-01 | 1 | 0 | 1 | 1 | 1 | 4 |
| 268 | rs143370854 | 20 | 1.01e-01 | 0 | 0 | 0 | 1 | 0 | 1 |
| 233 | rs75138251 | 9 | 1.07e-01 | 0 | 0 | 1 | 0 | 0 | 1 |
| 83 | rs61775691 | 1 | 1.07e-01 | 0 | 0 | 0 | 0 | 1 | 1 |
| 54 | rs111915634 | 4 | 1.07e-01 | 0 | 0 | 1 | 1 | 0 | 2 |
| 199 | rs79188580 | 8 | 1.09e-01 | 0 | 1 | 0 | 0 | 0 | 1 |
| 115 | rs59303138 | 3 | 1.10e-01 | 0 | 1 | 0 | 0 | 0 | 1 |

**Table S11:** Selected SNPs, fetal genome.
| Rank | SNP | Chromosome | p-value | Fold |  |  |  |  | Total |
| --- | --- | --- | --- | --- | --- | --- | --- | --- | --- |
|  |  |  |  | 0 | 1 | 2 | 3 | 4 |  |
| 87 | rs72846993 | 2 | 1.13e-01 | 0 | 1 | 0 | 0 | 0 | 1 |
| 121 | rs528779602 | 3 | 1.16e-01 | 0 | 1 | 0 | 0 | 0 | 1 |
| 90 | rs80201544 | 1 | 1.18e-01 | 0 | 0 | 0 | 0 | 1 | 1 |
| 256 | rs6011421 | 20 | 1.18e-01 | 0 | 1 | 0 | 0 | 0 | 1 |
| 203 | rs8192653 | 8 | 1.19e-01 | 1 | 0 | 0 | 0 | 0 | 1 |
| 86 | rs190247930 | 1 | 1.19e-01 | 0 | 0 | 0 | 1 | 0 | 1 |
| 68 | rs145427459 | 4 | 1.21e-01 | 0 | 0 | 1 | 0 | 1 | 2 |
| 217 | rs76230666 | 17 | 1.22e-01 | 1 | 0 | 0 | 1 | 0 | 2 |
| 225 | rs77691766 | 8 | 1.22e-01 | 0 | 0 | 0 | 0 | 1 | 1 |
| 116 | rs117946652 | 16 | 1.25e-01 | 1 | 0 | 1 | 1 | 0 | 3 |
| 269 | rs2092155 | 23 | 1.26e-01 | 0 | 1 | 0 | 0 | 1 | 2 |
| 260 | rs143117168 | 17 | 1.33e-01 | 1 | 0 | 0 | 0 | 0 | 1 |
| 97 | rs62233001 | 3 | 1.33e-01 | 0 | 0 | 1 | 0 | 0 | 1 |
| 0 | rs71638874 | 1 | 1.36e-01 | 1 | 1 | 1 | 1 | 1 | 5 |
| 123 | rs575089268 | 3 | 1.43e-01 | 0 | 1 | 0 | 0 | 0 | 1 |
| 160 | rs573055718 | 3 | 1.45e-01 | 0 | 0 | 0 | 0 | 1 | 1 |

**Table S11:** Selected SNPs, fetal genome.
| Rank | SNP | Chromosome | p-value | Fold |  |  |  |  | Total |
| --- | --- | --- | --- | --- | --- | --- | --- | --- | --- |
|  |  |  |  | 0 | 1 | 2 | 3 | 4 |  |
| 177 | rs72804004 | 5 | 1.47e-01 | 0 | 0 | 1 | 0 | 0 | 1 |
| 181 | rs9275733 | 6 | 1.48e-01 | 0 | 0 | 0 | 1 | 0 | 1 |
| 184 | rs538221772 | 8 | 1.52e-01 | 0 | 1 | 0 | 0 | 0 | 1 |
| 148 | rs79351607 | 3 | 1.54e-01 | 0 | 0 | 0 | 0 | 1 | 1 |
| 66 | rs1133684 | 5 | 1.55e-01 | 0 | 1 | 1 | 0 | 0 | 2 |
| 161 | rs140035409 | 3 | 1.56e-01 | 1 | 0 | 0 | 0 | 0 | 1 |
| 95 | rs11695051 | 2 | 1.57e-01 | 0 | 0 | 0 | 1 | 0 | 1 |
| 32 | rs115810594 | 2 | 1.57e-01 | 1 | 0 | 1 | 0 | 0 | 2 |
| 67 | rs35323270 | 5 | 1.59e-01 | 0 | 0 | 0 | 1 | 1 | 2 |
| 276 | rs11697094 | 20 | 1.62e-01 | 0 | 0 | 1 | 0 | 0 | 1 |
| 193 | rs76739100 | 6 | 1.63e-01 | 0 | 0 | 1 | 0 | 0 | 1 |
| 48 | rs72714356 | 8 | 1.65e-01 | 0 | 1 | 1 | 1 | 1 | 4 |
| 135 | rs116163066 | 3 | 1.67e-01 | 0 | 0 | 0 | 0 | 1 | 1 |
| 129 | rs4607069 | 3 | 1.68e-01 | 0 | 1 | 0 | 0 | 0 | 1 |
| 239 | rs117565066 | 16 | 1.72e-01 | 0 | 0 | 0 | 1 | 0 | 1 |
| 273 | rs144327896 | 23 | 1.73e-01 | 0 | 1 | 0 | 0 | 0 | 1 |

**Table S11:** Selected SNPs, fetal genome.
| Rank | SNP | Chromosome | p-value | Fold |  |  |  |  | Total |
| --- | --- | --- | --- | --- | --- | --- | --- | --- | --- |
|  |  |  |  | 0 | 1 | 2 | 3 | 4 |  |
| 270 | rs140940759 | 23 | 1.77e-01 | 0 | 1 | 0 | 0 | 0 | 1 |
| 47 | rs113129860 | 3 | 1.80e-01 | 1 | 0 | 0 | 1 | 0 | 2 |
| 190 | rs72850156 | 6 | 1.84e-01 | 0 | 0 | 0 | 0 | 1 | 1 |
| 218 | rs78336472 | 17 | 1.89e-01 | 1 | 1 | 0 | 0 | 0 | 2 |
| 197 | rs187467733 | 8 | 1.93e-01 | 0 | 0 | 0 | 1 | 0 | 1 |
| 185 | rs59922082 | 6 | 1.95e-01 | 0 | 0 | 0 | 1 | 0 | 1 |
| 162 | rs35526928 | 5 | 2.01e-01 | 0 | 1 | 0 | 0 | 0 | 1 |
| 20 | rs34744948 | 6 | 2.01e-01 | 1 | 1 | 0 | 1 | 1 | 4 |
| 234 | rs117133832 | 17 | 2.02e-01 | 1 | 1 | 0 | 0 | 0 | 2 |
| 164 | rs74542471 | 4 | 2.02e-01 | 0 | 0 | 1 | 0 | 0 | 1 |
| 183 | rs143270285 | 17 | 2.03e-01 | 1 | 0 | 1 | 1 | 0 | 3 |
| 118 | rs80157733 | 3 | 2.10e-01 | 0 | 0 | 0 | 0 | 1 | 1 |
| 221 | rs149713782 | 9 | 2.14e-01 | 1 | 0 | 0 | 0 | 0 | 1 |
| 12 | rs146272610 | 3 | 2.16e-01 | 1 | 0 | 1 | 1 | 0 | 3 |
| 29 | rs80212941 | 1 | 2.19e-01 | 0 | 1 | 0 | 1 | 0 | 2 |
| 103 | rs113664570 | 3 | 2.20e-01 | 0 | 0 | 1 | 0 | 0 | 1 |

**Table S11:** Selected SNPs, fetal genome.
| Rank | SNP | Chromosome | p-value | Fold |  |  |  |  | Total |
| --- | --- | --- | --- | --- | --- | --- | --- | --- | --- |
|  |  |  |  | 0 | 1 | 2 | 3 | 4 |  |
| 119 | rs2332238 | 3 | 2.26e-01 | 1 | 0 | 0 | 0 | 0 | 1 |
| 219 | rs138441221 | 9 | 2.28e-01 | 0 | 0 | 0 | 1 | 0 | 1 |
| 30 | rs140224603 | 1 | 2.34e-01 | 1 | 1 | 0 | 0 | 0 | 2 |
| 169 | rs111567122 | 4 | 2.39e-01 | 1 | 0 | 0 | 0 | 0 | 1 |
| 8 | rs77217171 | 4 | 2.40e-01 | 1 | 1 | 1 | 1 | 1 | 5 |
| 202 | rs72622848 | 8 | 2.40e-01 | 0 | 0 | 1 | 0 | 0 | 1 |
| 228 | rs37846 | 16 | 2.41e-01 | 0 | 1 | 0 | 0 | 0 | 1 |
| 210 | rs62527876 | 8 | 2.42e-01 | 0 | 0 | 0 | 1 | 0 | 1 |
| 281 | rs5910985 | 23 | 2.53e-01 | 0 | 0 | 0 | 1 | 0 | 1 |
| 42 | rs9267484 | 6 | 2.57e-01 | 0 | 0 | 1 | 1 | 1 | 3 |
| 73 | rs72714697 | 8 | 2.69e-01 | 1 | 0 | 1 | 1 | 0 | 3 |
| 101 | rs146923460 | 17 | 2.71e-01 | 1 | 1 | 1 | 1 | 1 | 5 |
| 34 | rs61106206 | 3 | 2.73e-01 | 1 | 0 | 1 | 0 | 0 | 2 |
| 98 | rs4852974 | 2 | 2.73e-01 | 0 | 0 | 0 | 0 | 1 | 1 |
| 51 | rs73231266 | 3 | 2.74e-01 | 1 | 0 | 0 | 1 | 0 | 2 |
| 255 | rs11078186 | 17 | 2.76e-01 | 0 | 0 | 0 | 1 | 0 | 1 |

**Table S11:** Selected SNPs, fetal genome.
| Rank | SNP | Chromosome | p-value | Fold |  |  |  |  | Total |
| --- | --- | --- | --- | --- | --- | --- | --- | --- | --- |
|  |  |  |  | 0 | 1 | 2 | 3 | 4 |  |
| 283 | rs149907900 | 23 | 2.80e-01 | 0 | 0 | 0 | 0 | 1 | 1 |
| 188 | rs117220763 | 6 | 2.80e-01 | 1 | 0 | 0 | 0 | 0 | 1 |
| 205 | rs3128935 | 6 | 2.81e-01 | 0 | 0 | 0 | 0 | 1 | 1 |
| 147 | rs139407759 | 4 | 2.91e-01 | 0 | 1 | 0 | 0 | 0 | 1 |
| 63 | rs116875306 | 16 | 3.00e-01 | 1 | 1 | 1 | 1 | 1 | 5 |
| 100 | rs12621351 | 2 | 3.02e-01 | 0 | 0 | 0 | 0 | 1 | 1 |
| 106 | rs76317024 | 9 | 3.05e-01 | 1 | 0 | 1 | 1 | 0 | 3 |
| 206 | rs67853213 | 17 | 3.15e-01 | 0 | 1 | 0 | 0 | 1 | 2 |
| 111 | rs184745845 | 3 | 3.26e-01 | 0 | 0 | 0 | 0 | 1 | 1 |
| 50 | rs73010237 | 3 | 3.29e-01 | 0 | 1 | 1 | 0 | 0 | 2 |
| 37 | rs4685205 | 3 | 3.31e-01 | 0 | 0 | 1 | 0 | 1 | 2 |
| 230 | rs72665204 | 8 | 3.34e-01 | 0 | 0 | 0 | 0 | 1 | 1 |
| 16 | rs17216347 | 3 | 3.35e-01 | 1 | 0 | 1 | 0 | 1 | 3 |
| 104 | rs150675350 | 16 | 3.37e-01 | 0 | 1 | 0 | 1 | 1 | 3 |
| 151 | rs151246320 | 8 | 3.41e-01 | 0 | 1 | 0 | 0 | 1 | 2 |
| 11 | rs62232959 | 3 | 3.46e-01 | 1 | 1 | 0 | 0 | 1 | 3 |

**Table S11:** Selected SNPs, fetal genome.
| Rank | SNP | Chromosome | p-value | Fold |  |  |  |  | Total |
| --- | --- | --- | --- | --- | --- | --- | --- | --- | --- |
|  |  |  |  | 0 | 1 | 2 | 3 | 4 |  |
| 216 | rs62574455 | 9 | 3.46e-01 | 0 | 1 | 0 | 0 | 0 | 1 |
| 284 | rs45584439 | 23 | 3.49e-01 | 0 | 1 | 0 | 0 | 0 | 1 |
| 19 | rs112214065 | 6 | 3.57e-01 | 1 | 0 | 1 | 1 | 1 | 4 |
| 240 | rs74656135 | 17 | 3.57e-01 | 0 | 0 | 1 | 0 | 1 | 2 |
| 139 | rs11926533 | 3 | 3.58e-01 | 1 | 0 | 0 | 0 | 0 | 1 |
| 186 | rs147255257 | 16 | 3.62e-01 | 1 | 0 | 0 | 1 | 0 | 2 |
| 6 | rs7650618 | 3 | 3.63e-01 | 1 | 1 | 1 | 1 | 0 | 4 |
| 74 | rs10733029 | 1 | 3.66e-01 | 1 | 0 | 0 | 0 | 0 | 1 |
| 275 | rs2001124 | 23 | 3.66e-01 | 1 | 0 | 1 | 0 | 0 | 2 |
| 15 | rs438475 | 6 | 3.69e-01 | 1 | 1 | 1 | 1 | 1 | 5 |
| 264 | rs141453072 | 23 | 3.70e-01 | 1 | 1 | 0 | 0 | 0 | 2 |
| 60 | rs116235998 | 4 | 3.72e-01 | 0 | 0 | 1 | 0 | 1 | 2 |
| 56 | rs114310352 | 6 | 3.81e-01 | 1 | 0 | 0 | 1 | 1 | 3 |
| 245 | rs35477639 | 17 | 3.86e-01 | 0 | 1 | 0 | 0 | 0 | 1 |
| 1 | rs116218025 | 1 | 3.89e-01 | 1 | 1 | 1 | 1 | 1 | 5 |
| 92 | rs111560165 | 1 | 3.94e-01 | 0 | 0 | 0 | 0 | 1 | 1 |

**Table S11:** Selected SNPs, fetal genome.
| Rank | SNP | Chromosome | p-value | Fold |  |  |  |  | Total |
| --- | --- | --- | --- | --- | --- | --- | --- | --- | --- |
|  |  |  |  | 0 | 1 | 2 | 3 | 4 |  |
| 187 | rs13249908 | 8 | 3.94e-01 | 0 | 1 | 0 | 0 | 0 | 1 |
| 53 | rs116172575 | 6 | 3.97e-01 | 1 | 1 | 0 | 0 | 1 | 3 |
| 158 | rs816547 | 3 | 4.00e-01 | 0 | 0 | 0 | 0 | 1 | 1 |
| 159 | rs72708267 | 4 | 4.01e-01 | 0 | 1 | 0 | 0 | 0 | 1 |
| 172 | rs193240735 | 4 | 4.05e-01 | 1 | 0 | 0 | 0 | 0 | 1 |
| 212 | rs73261904 | 17 | 4.08e-01 | 1 | 0 | 1 | 1 | 0 | 3 |
| 58 | rs73503978 | 16 | 4.13e-01 | 1 | 1 | 1 | 1 | 1 | 5 |
| 257 | rs5930433 | 23 | 4.16e-01 | 1 | 1 | 0 | 1 | 1 | 4 |
| 211 | rs77822078 | 17 | 4.23e-01 | 1 | 0 | 0 | 1 | 0 | 2 |
| 241 | rs149926327 | 17 | 4.31e-01 | 0 | 1 | 0 | 0 | 0 | 1 |
| 31 | rs140529843 | 1 | 4.43e-01 | 1 | 0 | 0 | 0 | 1 | 2 |
| 208 | rs149746636 | 16 | 4.46e-01 | 0 | 0 | 1 | 0 | 1 | 2 |
| 243 | rs78657521 | 17 | 4.56e-01 | 0 | 0 | 0 | 1 | 0 | 1 |
| 207 | rs111305015 | 17 | 4.67e-01 | 0 | 0 | 1 | 1 | 0 | 2 |
| 277 | rs191463305 | 23 | 4.75e-01 | 0 | 0 | 0 | 1 | 0 | 1 |
| 274 | rs142557034 | 23 | 4.78e-01 | 0 | 0 | 0 | 0 | 1 | 1 |

**Table S11:** Selected SNPs, fetal genome.
| Rank | SNP | Chromosome | p-value | Fold |  |  |  |  | Total |
| --- | --- | --- | --- | --- | --- | --- | --- | --- | --- |
|  |  |  |  | 0 | 1 | 2 | 3 | 4 |  |
| 105 | rs12490230 | 3 | 4.82e-01 | 0 | 0 | 1 | 0 | 0 | 1 |
| 138 | rs145930653 | 3 | 5.10e-01 | 0 | 0 | 1 | 0 | 0 | 1 |
| 22 | rs4683629 | 3 | 5.10e-01 | 0 | 0 | 1 | 1 | 1 | 3 |
| 85 | rs67874885 | 1 | 5.11e-01 | 0 | 1 | 0 | 0 | 0 | 1 |
| 272 | rs35183165 | 20 | 5.42e-01 | 0 | 0 | 0 | 1 | 0 | 1 |
| 142 | rs73252944 | 4 | 5.42e-01 | 0 | 0 | 0 | 1 | 0 | 1 |
| 213 | rs139078603 | 8 | 5.50e-01 | 1 | 0 | 0 | 0 | 0 | 1 |
| 279 | rs6010742 | 20 | 5.60e-01 | 0 | 0 | 1 | 0 | 0 | 1 |
| 109 | rs79133272 | 3 | 5.79e-01 | 1 | 0 | 0 | 0 | 0 | 1 |
| 122 | rs146005261 | 3 | 5.81e-01 | 0 | 0 | 0 | 1 | 0 | 1 |
| 189 | rs147137688 | 8 | 5.81e-01 | 0 | 1 | 0 | 0 | 0 | 1 |
| 156 | rs117362350 | 8 | 5.84e-01 | 0 | 1 | 1 | 0 | 0 | 2 |
| 141 | rs143940877 | 4 | 5.86e-01 | 0 | 1 | 0 | 0 | 0 | 1 |
| 28 | rs72678377 | 1 | 5.89e-01 | 1 | 1 | 0 | 0 | 0 | 2 |
| 209 | rs76221837 | 17 | 5.91e-01 | 0 | 1 | 1 | 0 | 0 | 2 |
| 52 | rs138072779 | 3 | 5.98e-01 | 1 | 0 | 1 | 0 | 0 | 2 |

**Table S11:** Selected SNPs, fetal genome.
| Rank | SNP | Chromosome | p-value | Fold |  |  |  |  | Total |
| --- | --- | --- | --- | --- | --- | --- | --- | --- | --- |
|  |  |  |  | 0 | 1 | 2 | 3 | 4 |  |
| 108 | rs73137746 | 3 | 6.05e-01 | 0 | 0 | 0 | 0 | 1 | 1 |
| 180 | rs17587604 | 6 | 6.32e-01 | 0 | 1 | 0 | 0 | 0 | 1 |
| 236 | rs113757163 | 17 | 6.38e-01 | 0 | 0 | 1 | 1 | 1 | 3 |
| 91 | rs11673787 | 2 | 6.41e-01 | 0 | 0 | 0 | 1 | 0 | 1 |
| 165 | rs74828088 | 8 | 6.53e-01 | 0 | 0 | 0 | 1 | 1 | 2 |
| 153 | rs12513307 | 4 | 6.62e-01 | 0 | 0 | 0 | 1 | 0 | 1 |
| 195 | rs28537590 | 8 | 6.64e-01 | 0 | 1 | 0 | 0 | 0 | 1 |
| 82 | rs150479064 | 1 | 6.65e-01 | 0 | 0 | 0 | 1 | 0 | 1 |
| 263 | rs189031310 | 20 | 6.68e-01 | 0 | 0 | 0 | 1 | 0 | 1 |
| 137 | rs147735769 | 3 | 6.72e-01 | 0 | 1 | 0 | 0 | 0 | 1 |
| 222 | rs2181034 | 9 | 6.72e-01 | 0 | 0 | 0 | 1 | 0 | 1 |
| 132 | rs149316678 | 3 | 6.78e-01 | 0 | 0 | 0 | 0 | 1 | 1 |
| 259 | rs77698168 | 17 | 6.78e-01 | 0 | 0 | 0 | 0 | 1 | 1 |
| 4 | rs151269448 | 3 | 6.79e-01 | 1 | 1 | 1 | 1 | 1 | 5 |
| 174 | rs141491495 | 9 | 6.81e-01 | 1 | 0 | 0 | 0 | 1 | 2 |
| 249 | rs2024088 | 17 | 6.88e-01 | 0 | 1 | 0 | 0 | 0 | 1 |

**Table S11:** Selected SNPs, fetal genome.
| Rank | SNP | Chromosome | p-value | Fold |  |  |  |  | Total |
| --- | --- | --- | --- | --- | --- | --- | --- | --- | --- |
|  |  |  |  | 0 | 1 | 2 | 3 | 4 |  |
| 78 | rs115022189 | 1 | 6.97e-01 | 1 | 0 | 0 | 0 | 0 | 1 |
| 200 | rs79350445 | 17 | 7.02e-01 | 1 | 0 | 0 | 1 | 1 | 3 |
| 120 | rs117180124 | 8 | 7.15e-01 | 0 | 1 | 0 | 1 | 0 | 2 |
| 285 | rs139428685 | 23 | 7.20e-01 | 0 | 1 | 0 | 0 | 0 | 1 |
| 150 | rs1038115 | 4 | 7.20e-01 | 0 | 0 | 0 | 1 | 0 | 1 |
| 94 | rs116382491 | 2 | 7.21e-01 | 0 | 1 | 0 | 0 | 0 | 1 |
| 246 | rs11649484 | 16 | 7.38e-01 | 0 | 0 | 0 | 0 | 1 | 1 |
| 124 | rs55764682 | 3 | 7.41e-01 | 1 | 0 | 0 | 0 | 0 | 1 |
| 70 | rs77419727 | 4 | 7.42e-01 | 1 | 0 | 1 | 0 | 0 | 2 |
| 223 | rs9915627 | 17 | 7.43e-01 | 1 | 0 | 1 | 1 | 0 | 3 |
| 215 | rs117775004 | 8 | 7.49e-01 | 0 | 0 | 1 | 0 | 0 | 1 |
| 128 | rs145321980 | 3 | 7.56e-01 | 1 | 0 | 0 | 0 | 0 | 1 |
| 171 | rs116152825 | 4 | 7.58e-01 | 0 | 0 | 1 | 0 | 0 | 1 |
| 238 | rs758434 | 17 | 7.72e-01 | 0 | 1 | 0 | 0 | 0 | 1 |
| 253 | rs12939398 | 17 | 7.74e-01 | 0 | 0 | 0 | 0 | 1 | 1 |
| 25 | rs183708312 | 1 | 7.93e-01 | 0 | 1 | 0 | 0 | 1 | 2 |

**Table S11:** Selected SNPs, fetal genome.
| Rank | SNP | Chromosome | p-value | Fold |  |  |  |  | Total |
| --- | --- | --- | --- | --- | --- | --- | --- | --- | --- |
|  |  |  |  | 0 | 1 | 2 | 3 | 4 |  |
| 266 | rs185781576 | 20 | 8.20e-01 | 1 | 0 | 0 | 0 | 0 | 1 |
| 265 | rs113698135 | 17 | 8.22e-01 | 0 | 0 | 0 | 0 | 1 | 1 |
| 46 | rs75543533 | 3 | 8.29e-01 | 0 | 0 | 1 | 0 | 1 | 2 |
| 26 | rs76266717 | 1 | 8.50e-01 | 0 | 0 | 1 | 1 | 0 | 2 |
| 146 | rs10016898 | 4 | 8.52e-01 | 0 | 0 | 0 | 1 | 0 | 1 |
| 2 | rs41302778 | 1 | 8.94e-01 | 1 | 1 | 1 | 1 | 1 | 5 |
| 131 | rs74669781 | 3 | 9.06e-01 | 0 | 0 | 1 | 0 | 0 | 1 |
| 69 | rs150970446 | 16 | 9.27e-01 | 1 | 1 | 1 | 1 | 1 | 5 |
| 75 | rs79822094 | 1 | 9.32e-01 | 0 | 0 | 0 | 1 | 0 | 1 |
| 14 | rs76416590 | 3 | 9.44e-01 | 1 | 0 | 1 | 0 | 1 | 3 |
| 250 | rs191178303 | 17 | 9.68e-01 | 0 | 0 | 1 | 0 | 0 | 1 |

